# Integrating Whole Genome and Transcriptome Sequencing to Characterize the Genetic Architecture of Isoform Variation and its Implications for Health and Disease

**DOI:** 10.1101/2024.12.04.24318434

**Authors:** Chunyu Liu, Roby Joehanes, Jiantao Ma, Jiuyong Xie, Jian Yang, Mengyao Wang, Tianxiao Huan, Shih-Jen Hwang, Jia Wen, Quan Sun, Demirkale Y. Cumhur, Nancy L. Heard-Costa, Peter Orchard, April P. Carson, Laura M. Raffield, Alexander Reiner, Yun Li, George O’Connor, Joanne M. Murabito, Peter Munson, Daniel Levy

## Abstract

We created a comprehensive whole blood splice variation quantitative trait locus (sQTL) resource by analyzing isoform expression ratio (isoform-to-gene) in Framingham Heart Study (FHS) participants (discovery: n=2,622; validation: n=1,094) with whole genome (WGS) and transcriptome sequencing (RNA-seq) data. External replication was conducted using WGS and RNA-seq from the Jackson Heart Study (JHS, n=1,020). We identified over 3.5 million *cis*-sQTL-isoform pairs (*p*<5e-8), comprising 1,176,624 *cis*-sQTL variants and 10,883 isoform transcripts from 4,971 sGenes, with significant change in isoform-to-gene ratio due to allelic variation. We validated 61% of these pairs in the FHS validation sample (*p*<1e-4). External validation (*p*<1e-4) in JHS for the top 10,000 and 100,000 most significant *cis*-sQTL-isoform pairs was 88% and 69%, respectively, while overall pairs validated at 23%. For 20% of *cis*-sQTLs in the FHS discovery sample, allelic variation did not significantly correlate with overall gene expression. sQTLs are enriched in splice donor and acceptor sites, as well as in GWAS SNPs, methylation QTLs, and protein QTLs. We detailed several sentinel *cis*-sQTLs influencing alternative splicing, with potential causal effects on cardiovascular disease risk. Notably, rs12898397 (T>C) affects splicing of *ULK3*, lowering levels of the full-length transcript ENST00000440863.7 and increasing levels of the truncated transcript ENST00000569437.5, encoding proteins of different lengths. Mendelian randomization analysis demonstrated that a lower ratio of the full-length isoform is causally associated with lower diastolic blood pressure and reduced lymphocyte percentages. This sQTL resource provides valuable insights into how transcriptomic variation may influence health outcomes.

## INTRODUCTION

Transcriptomic isoform variation generates different messenger RNAs (mRNAs) from the same pre-mRNA, greatly expanding proteomic diversity from the ∼20,000 protein-coding genes in the human genome.^1^ One mechanism for transcriptomic isoform variation is alternative splicing (AS), which joins exons in different combinations to produce mature RNA isoforms.^2^ Isoform variation may be a direct cause or a consequence of disease.^3,4^ Owing to advances in sequencing technologies, recent studies have revealed that up to 95% of multi-exon genes undergo AS in humans.^1^

While it is a dynamic process, isoform variation is under tight regulation. Many variables, such as *cis*- or *trans*-regulatory elements and DNA methylation patterns regulate isoform variation and AS.^5–7^ Several previous studies have provided crucial insights into the genetic regulation of transcriptomic isoform variation, contributing to our understanding of the genetic architecture underlying complex diseases.^8–17^ Two studies used whole-genome (WGS) and whole-transcriptome sequencing (RNA-seq) from the Genotype-Tissue Expression (GTEx) consortium to identify single nucleotide polymorphisms (SNPs) affecting splice variation throughout the human genome across multiple tissues.^14,18^ They found that expression levels of nearly all expressed genes (eGenes) are subject to genetic regulation.^18^ In addition, SNPs that influence splicing (splicing quantitative trait loci or sQTLs) can exert a more pronounced impact on complex traits compared to those affecting gene expression levels (expression quantitative trait loci or eQTLs) alone. ^14^ The replication rate for sQTLs identified in these two studies, however, was low, possibly due to methodological differences in isoform characterization.^14,18^

A comprehensive analysis of the association of sQTLs with isoform variation in whole blood and the implications of these findings for human health and disease is limited. To that end, we conducted sQTL analyses using WGS and RNA-seq data from whole-blood derived DNA and RNA obtained from 3716 Framingham Heart Study (FHS) participants. We conducted external validation using WGS and RNA-seq data (in peripheral blood mononuclear cells (PBMCs)) from the Jackson Heart Study (JHS).^19^ The overarching aims of our study were to create a comprehensive sQTL resource and to demonstrate the relations of isoform variation to several human traits (**Figure 1**). We distinguished sQTLs with little or no effect on overall gene-level expression from sQTLs that also function as eQTLs. We identified sQTLs that coincide with trait-associated variants reported in genome-wide association studies (GWAS) and used Mendelian randomization (MR) to infer causal relations of isoform variation to disease-related traits, with a focus on cardiovascular disease (CVD) risk factors and white blood cell (WBC) composition. In addition, we evaluated for enrichment of sQTLs in other types of QTLs, including those associated with DNA methylation (mQTLs) and plasma protein levels (pQTLs). We provide three examples of sQTLs and their corresponding transcripts that demonstrate how our sQTL resource can identify putatively causal genetic variants impacting human diseases through regulatory effects on transcriptomic variation.

**Figure 1.**
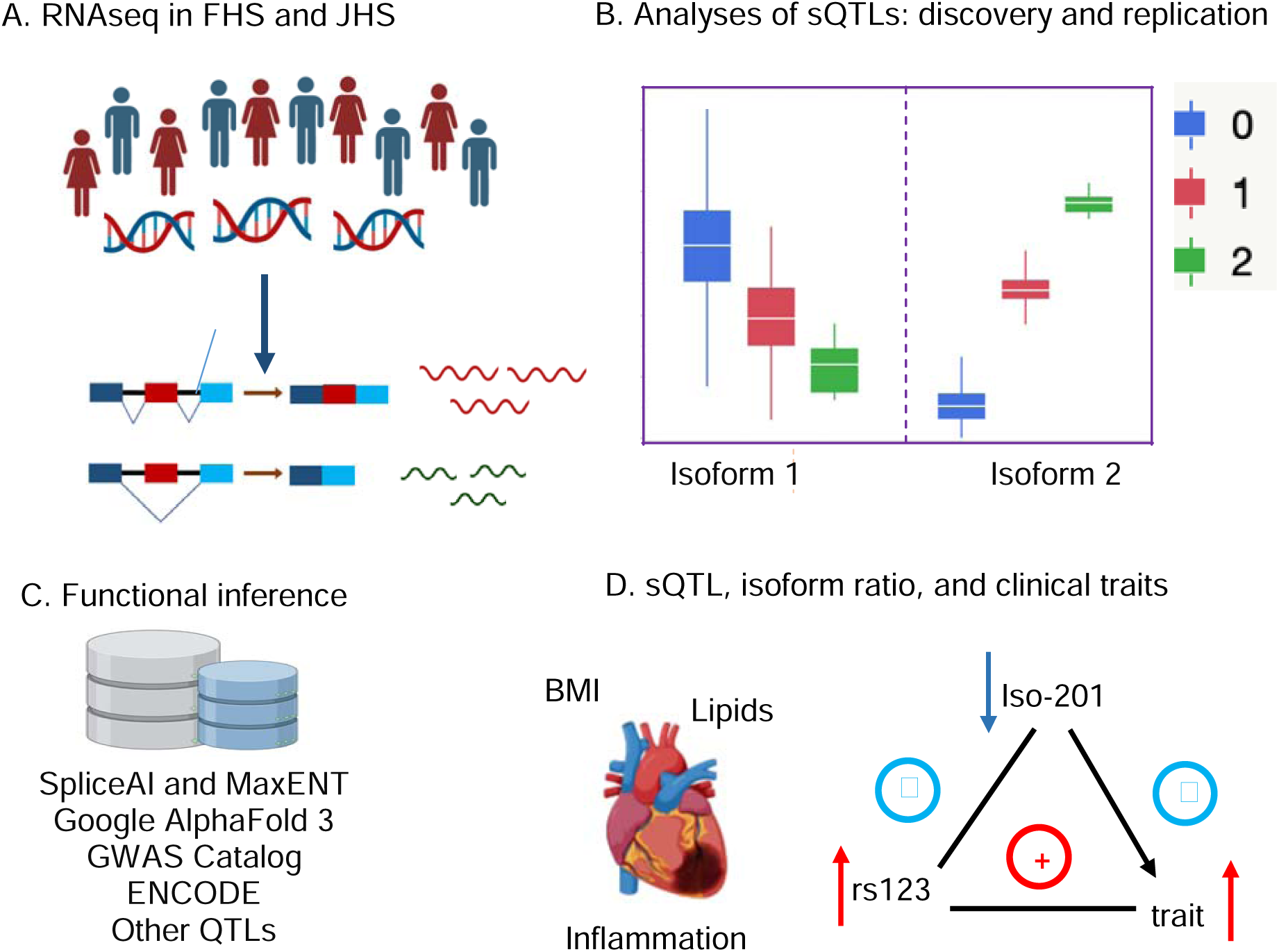
Schematic Outline. (A) RNA sequencing in Framingham Heart Study (FHS). Stored blood samples from a community-based population provide complete genome sequence and expression analysis of multiple, alternatively spliced transcripts for thousands of genes. (B) Analyses of eQTLs and sQTLs. Overall gene expression and expression ratio of each transcript isoform may depend on nearby genomic variants. The number of alleles of a particular variant (0, 1, or 2) present may be associated with low, mid-range, or high expression of an isoform, resulting from a particular pattern of mRNA splicing. (C) Functional Inference. The consequences of genetically determined alternative splicing are assessed with tools such as Gene Set Enrichment Analysis and by reference to databases such as the Genome-Wide Association Study (GWAS) Catalog. (D) sQTL, isoform ratio and clinical traits: regression and MR analysis. The cardiovascular and hematological implications of common genetic variants and alternative splicing are assessed by combining our findings with clinical measurements in the FHS, and GWAS catalog, and using Mendelian randomization, establish the causal connection between the genetic variant, alternative splicing, and propensity to manifest the trait or disease.

## METHODS

### Study participants

We included 3,716 self-identified White participants in the FHS Offspring and Third Generation cohorts for discovery and internal validation (**Table 1**).^20,21^ Blood samples for RNA-seq were collected at the ninth examination cycle (2011–2014) of the Offspring cohort participants and the second examination cycle (2008–2011) of the Third Generation cohort participants. For replication, we included self-identified 1,020 African American participants in the JHS^22^ (**Table 1**). Blood samples for RNA-seq were collected during examination cycle one (2000 - 2004). In both JHS and FHS, we did not exclude any participants based on genetic similarity to reference panels such as 1000G. Protocols for the examination of participants and the collection of genetic materials were approved by the Institutional Review Board at Boston University Medical Center for FHS and the University of Mississippi Medical Center (UMMC) for JHS. All participants provided written, informed consent for genetic studies. All research was performed in accordance with relevant guidelines/regulations.

**Table 1.**
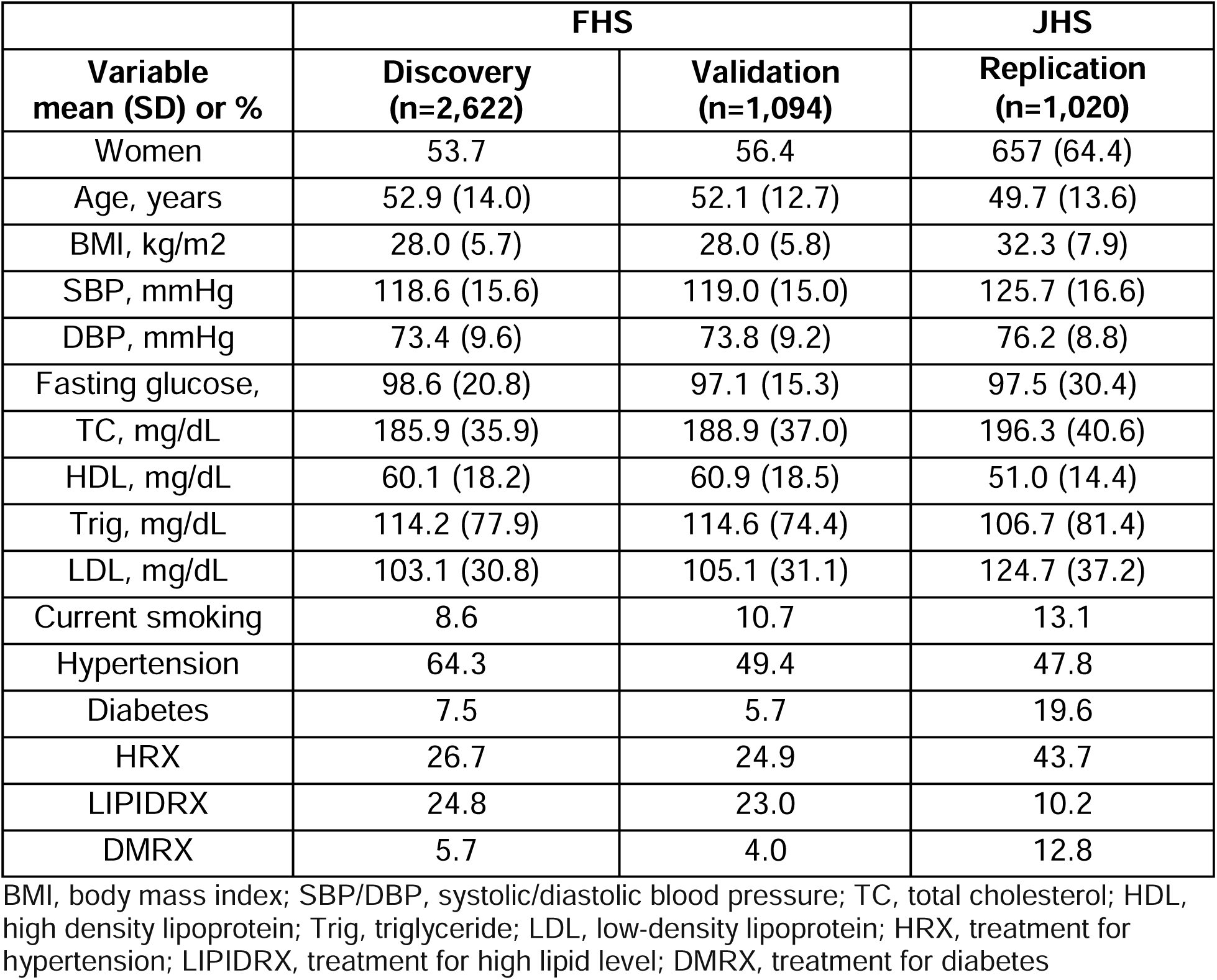
Participant characteristics of the discovery and validation samples in the Framingham Heart Study.

### Isolation of RNA from whole blood

Isolation of RNA from peripheral whole blood was described previously.^23,24^ PAXgene™ tubes (PreAnalytiX, Hombrechtikon, Switzerland) were used to collect peripheral whole blood samples (2.5 mL) from the FHS participants. The collected blood samples were incubated at room temperature for 4 hours for RNA stabilization, and then stored at −80 °C until use. Tubes were allowed to thaw for 16 hours at room temperature before RNA extraction. A standard protocol was used to extract total RNA with a PAXgene Blood RNA Kit at the FHS Genetics Laboratory (FHS Third Generation cohort) and the TOPMed contract laboratory at Northwest Genomics Center (Offspring cohort). After centrifugation and washing, white blood cell pellets were collected and lysed in guanidinium-containing buffer. The extracted RNA was evaluated for quality by determining absorbance readings at 260 and 280 nm using a NanoDrop ND-1000 UV spectrophotometer. The integrity of total RNA was determined with the Agilent Bioanalyzer 2100 microfluidic electrophoresis (Nano Assay and the Caliper LabChip system).

In JHS, PBMC samples from 1012 samples were used for RNA extraction at the University of Washington Northwest Genomics Center (NWGC). Detailed information for library construction and processing was previously described.^19^ In brief, total RNA was scaled to 7.5ng/ul (total volume of 50ul) on the Perkin Elmer Janus II Workstation to construct library. The TruSeq Stranded mRNA kit (Illumina, cat#RS-122-2103) was used for poly-A selection and cDNA synthesis were performed. Successful libraries were normalized and pooled before sequencing.

### Whole genome sequencing

Details on DNA extraction, processing, and identification of variants were previously described.^19^ In brief, DNA extracted from whole blood (in FHS) or PBMC (in JHS) underwent whole genome sequencing (WGS) at TOPMed contract sequencing centers. The FHS samples were sequenced at the Broad Institute while the JHS samples were sequenced at the University of Washington. Consistent sequencing and data processing criteria were used across all centers, with DNA sequence alignment to human genome build GRCh38. The resulting BAM files were sent to TOPMed’s Informatics Research Center (IRC) for re-alignment and standardized BAM file generation. This study used WGS data from Freeze 10a which included approximately 208 million SNPs across TOPMed cohorts.

### RNA sequencing and processing

In both FHS and JHS, RNA sequencing was conducted at the University of Washington Northwest Genomics Center, an NHLBI TOPMed program reference laboratory, following a standard protocol.^25^ Data processing was conducted by the University of Washington laboratory (**Supplemental Figure 1**). Base call generation was performed on the NovaSeq 6000 instrument (RTA 3.1.5), followed by the conversion of demultiplexed, unaligned BAM files to FASTQ format using SamTools bam2fq (v1.4). Sequence and base quality were assessed with the FASTX-toolkit (v0.0.13), and sequence alignment to GRCh38 with reference transcriptome GENCODE release 30 was conducted using STAR. FastQC (version 0.11.9) provided initial quality control metrics. A genome index for mapping was built by STAR to generate BAM files at the gene and transcript levels based on the GRCh38 reference build. The TOPMed RNA-seq pipeline utilized RNA-SeQC ^26^ to derive standard quality control metrics from aligned reads.

### Calculation of isoform ratio

Transcript-level expression of an isoform was quantified as reads per kilobase of transcript per million mapped reads (RPKM), a normalization measure that considers the gene length in quantifying the expression level of a transcript. The trimmed means of M-values (TMM)^27^ were derived from the FPKM values using the edgeR package.^26,28^ The TMM method estimates scale factors between samples and estimates relative RNA production levels from RNA-seq data. The isoform expression ratio was calculated as the transcript level of an isoform (TMM value) divided by the sum of the levels of all isoforms (the sum of the TMM values) of its parent gene. The transcript isoform ratio was considered as missing if the overall expression level of its gene was zero.^15^

### Association of genetic variants with isoform ratios

We performed linear regression analyses of genome-wide SNPs having minor allele frequencies (MAFs) ≥ 0.01 and Hardy-Weinburg Equilibrium (HWE) *p* ≥ 1e-10 with isoform ratios for transcripts having the population median TMM ≥ 2 and fewer than 20% missing values. The dosage (0, 1 or 2) of the alternative allele of each SNP was used as a continuous independent variable (additive model) and the isoform ratio was used as the dependent variable. We sought to minimize confounding by adjusting the isoform ratios by several covariates as follows.^29^ We adjusted for five principal components (PCs) computed from genotype profiles in the FHS to account for population stratification. We also adjusted for 15 PCs computed from the transcriptome profile to account for unknown confounders that may affect gene expression. Furthermore, we adjusted for family relationships by including a pedigree correlation structure as a random effect in the regression mode. We explored additional technical covariates, including year of blood collection, batch (sequencing machine and time, plate and well), and RNA concentration^29^ but these covariates were not included in the final models since they explained <1% of variance in isoform ratios. We employed the NIH-supported STRIDES cloud computing infrastructure for all association analyses. We used a Graphical Processing Unit (GPU)-based program ^24^ for all computations. We stored effect sizes, standard error, partial R-squared, and p-values for SNP-isoform pairs with *p*<1e-4, to facilitate future meta-analysis and lookups by the broad scientific community.

We identified significant isoform-ratio QTLs; for simplicity, we refer to them as sQTLs and their target genes as sGenes throughout. A *cis*-sQTL was defined as a polymorphism falling within 1 Mb of the transcription start site (TSS) of the isoform. A *trans*-sQTL refers to a polymorphism that is beyond 1 Mb from the TSS of its isoform or residing on the different chromosomes. We used *p*<5×10^−8^ to denote a significant *cis*-sQTL and *p*<1.5×10^−13^ (∼ 0.05/(25,642*12,887,093)) to define a significant *trans*-sQTL.

### Internal validation

Association analyses were conducted in two stages, a discovery stage (n=2,622) and a validation stage (n=1,094) (**Table 1**). A significant sQTL-transcript pair was considered validated if that pair received *p*<1e-4 (for both cis- and trans-pairs) and had consistent effect sign in the validation data.

### External replication

The JHS conducted RNA-seq for 1,022 African-American participants who also had WGS.^30^ We analyzed the JHS data using the isoform ratio method in 1,020 JHS participants after removing two participants with possible sample identification problems. Owing to differences in sample size, tissue (whole blood versus PBMCs), mRNA isolation techniques, and differences in study design (including but not limited to self-reported race, region, and recruitment criteria and methods), certain isoforms detected in the FHS samples (median TMM < 2, missingness < 20%) were not identified in the JHS samples. Similarly, some SNPs that met the inclusion criteria (MAF > 0.01, p-H-W < 1e-10) in FHS were not available in JHS based on these same filtering criteria. Therefore, the replication rate was calculated only for the sQTL-isoform pairs that were significant in FHS and met the inclusion criteria in JHS (“eligible pairs”). The replication rate was calculated as the ratio of the number of *cis-* or *trans*-sQTL-isoform pairs with p < 1e-4 and consistent effects (i.e., the same directionality for beta estimates) in both cohorts to the number of the significant sQTL-isoform pairs that met the inclusion criteria in JHS.

We also highlighted replication rates for the 10,000 and 100,000 most significant sQTL-isoform pairs identified in FHS using the same definition. We compared the MAFs, variance of isoform ratio explained (partial R^2^), and effect sizes of the sentinel sQTLs (of isoforms) in the FHS to those in the JHS. Although GTEx has produced an extensive resource for sQTL analysis,^13,14^ we conducted only limited replication analysis using the GTEx data owing to significant methodological differences between the FHS and GTEx approaches in identifying sQTLs (**Supplemental materials**).

### Comparison of functional relevance of *cis*-sQTLs and *cis*-eQTLs

We conducted several analyses to explore the functional relevance of *cis*-sQTLs in comparison to *cis*-eQTLs^23^ in the same participants. We compared the number of genes containing *cis*-sQTLs (*cis*-sGenes) to those containing *cis*-eQTLs (*cis*-eGenes). We also compared the *cis*-sQTLs and *cis*-eQTLs regarding their distance from their respective TSSs. Additionally, we evaluated the variance explained by *cis*-sQTLs compared to that explained by *cis*-eQTLs.

We assessed the enrichment of sentinel *cis*-sQTLs and *cis*-eQTLs in relevant genomic features, including splice donor/acceptor sites, intron/exon boundaries, and eCLIP peaks for binding sites of 203 RNA-binding proteins (RBPs) from ENCODE.^31,32^ To reduce bias, we generated 1,000 sets, consisting of randomly selected SNPs with MAF ≥1% and Hardy-Weinberg equilibrium p ≥1e-10, one for each cis-sQTL or cis-eQTL, matched based on similar MAF and distance to the transcription start site (TSS) for the respective *cis*-sQTLs or *cis*-eQTLs, using the following bin boundaries: MAF bins at 0.01, 0.05, 0.1, 0.2, 0.3, and 0.5, and TSS distance bins at 0kb, 1kb, 10kb, 100kb, and 1Mb. Enrichment of *cis*-sQTLs and *cis*-eQTLs in each feature was calculated as follows:

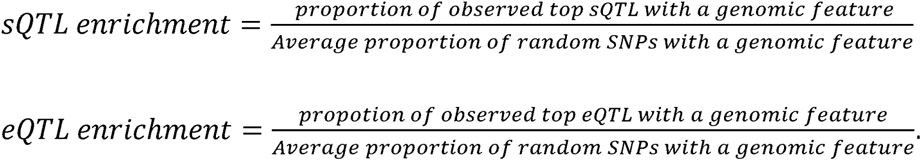

### Enrichment analysis of sQTLs with GWAS, mQTLs, and pQTLs

#### Overlap of sQTLs with GWAS Catalog SNPs

We selected GWAS Catalog SNPs^33^ using two criteria: 1) a significance threshold of *p*<5e-8, and 2) a sample size threshold reported with both discovery and replication sample sizes ≥5000, or the discovery sample size ≥10,000. Replicated sQTLs were those in the sQTL-isoform pairs with *p*<5e-8 in the discovery sample, and *p*<1e-4 in the validation and replication sample. We then matched the selected GWAS Catalog SNPs with the identified sQTLs.

#### Enrichment analyses using Fisher’s exact test

We evaluated the null hypothesis that sQTLs are independent of (i.e., not enriched with) 1) GWAS SNPs, 2) genetic variants associated with DNA methylation (mQTLs), or 3) protein (pQTLs). The mQTL data were obtained from GoDMC from more than 30,000 study participants with genetic and DNAm data from an international collaboration of human epidemiological studies.^34^ The pQTL database was obtained from a recent report that characterized the genetic basis of the plasma proteomic profiles in 54,219 UK Biobank participants.^35^ The total number of SNPs was 12,887,093 SNPs (MAF ≥1% and Hardy-Weinberg equilibrium *p* ≥1e-10) across the genome. For example, we created a 2 x 2 table that includes the number of overlapping SNPs between GWAS SNPs and sQTLs, the number of SNPs in GWAS SNPs but not in sQTLs, the number of SNPs of sQTLs but not in GWAS SNPs, and remaining SNPs using 12,887,093 as the reference. We conducted the Fisher’s exact test to determine the independence of sQTLs from GWAS SNPs, or mQTLs, or pQTLs.

### Prediction of SNP effects on alternative splicing

We used two methods, SpliceAI^36^ and MaxENT,^37^ to predict the SNP effects for selected sQTLs on transcriptomic splice variation. SpliceAI uses deep a neural network to identify splicing and helps predict non-coding cryptic splice variants in the human genome.^36^ MaxENT refers to maximum entropy models that are specified by changing a set of constraints to discriminate human DNA 5’GT (i.e., donor) and 3’AG (i.e., acceptor) splice sites from non-functional decoys.^37^

### Prediction of protein structures by Google AlphaFold 3.0

As an example of how genotype variants might affect protein structure, we used Google AlphaFold 3.0 to predict protein structures^38,39^ for two of the protein coding isoforms of *ULK3* (unc-51 like kinase 3) associated with genotype rs12898397 (T>C) (**Supplemental Materials**). The sQTL variant and the associated variation of protein sequence are in the COOH-terminal region of ULK3 (NM_001411082.1). To validate the structure predictions, we first aligned them with the crystal structure of the final MIT domain of ULK3, 4WZX from the Protein Data Bank^40^ using the PyMOL Molecular Graphics System (Version 3.0.4, Schrödinger, LLC).^38^ Next we compared the predicted structures in the COOH-terminal regions surrounding the consequent changes in protein sequence. The overlay of these two predicted structures was visualized in PyMOL. The structural differences were quantified using root-mean-square deviation (RMSD) values. An RMSD of 1.0 angstrom or lower indicates nearly identical conformations, while RMSD of 3.0 or higher suggests significant structural differences.^41^

### Mendelian randomization (MR) analysis

To illustrate the utility of our sQTL resource, we conducted MR analysis to infer causality between the isoform ratios (as the exposure) of the respective transcripts for selected isoforms that harbor significant *cis*-sQTLs and CVD factors and blood composition traits (as the outcome). *cis*-sQTLs served as the instrumental variables (IVs). We used the MR-base^42^ software to conduct the MR analysis in conjunction with summary statistics of SNPs from a large GWAS database^33^ (**Supplemental Table 1**). To minimize bias, we selected SNPs in MR analysis based on their linkage disequilibrium (LD) r^2^ < 0.001.^42^ We applied the inverse variance weighted (INV) method to summarize results in MR analyses and used MR-Egger method to assess SNP horizontal pleiotropic effects.^42^

All statistical analyses were conducted using R (version 4.4.0). Throughout the manuscript, we reported small p-values as -logP, representing -log_10_ (p-value).

## RESULTS

We analyzed associations of isoform ratios for 25,642 isoform transcripts with 12,887,093 SNPs (see Methods). We identified *cis*-sQTLs (at *p* <5×10^−8^) and *trans*-sQTLs (at *p* <1.5×10^−13^) in the FHS discovery (n=2,622) and validation (n=1,094) samples (*p* <1e-4), followed by external replication in 1,020 JHS participants. We compared of *cis*-sQTLs and *cis*-eQTLs and analyses of enrichment of *cis*-sQTLs in GWAS Catalog SNPs,^33^ *cis*-mQTLs,^34^ and *cis*-pQTLs.^23^ We also provide examples of how *cis*-sQTLs can be used in two-sample MR analysis to infer putatively causal relation of isoform variation to CVD risk factors and whole blood WBC composition (**Figure 1**).

### Transcriptome-wide Identification of *cis*- and *trans*-sQTLs in the FHS

In the FHS discovery sample, we identified a total of 3,588,153 significant *cis*-sQTL-isoform pairs, comprising 1,176,624 distinct *cis*-sQTL variants and 10,883 distinct isoform transcripts from 4,971 distinct sGenes (**Table 2)**. Of these, 61% of the *cis* pairs were validated in the FHS validation sample. We also identified 328,997 significant *trans*-sQTL-isoform pairs, consisting of 131,804 distinct *trans*-sQTL variants and 1,054 distinct isoform transcripts from 590 distinct sGenes in the FHS discovery sample. Compared to *cis*-pairs, a larger proportion of trans-sQTL-isoform pairs (n=235,807, 72%) were validated in the FHS validation sample (**Table 2**). The sentinel *cis*-sQTL and *trans*-sQTLs identified in FHS were presented in **Supplemental Tables 2 and 3**.

**Table 2.** The number of sQTL findings in the FHS and JHS participants.

|  | FHS (n = 3,716) |  |  | JHS (n = 1,020) |  |  |
| --- | --- | --- | --- | --- | --- | --- |
|  | Discovery | Validation |  | Replication |  |  |
| | *cis*: $p < 5e-8$<br>*trans*: $p < 1.5e-13$ | $p < 1e-4$ | % | Eligible pairs | $p < 1e-4$ | % |
|  | **Cis-** |  |  |  |  |  |
| SNP-Isoform Pairs | 3,588,153 | 2,193,946 | 61.1 | 1,583,143 | 358,654 | 22.7% |
| Distinct SNPs | 1,176,624 | 867,743 | 73.7 | 671,730 | 213,050 | 31.7% |
| Distinct isoforms | 10,883 | 8,307 | 76.3 | 4,453 | 2,546 | 57.2% |
| Distinct genes | 4,971 | 4,173 | 83.9 | 2,808 | 1,713 | 61.0% |
|  | **Trans-** |  |  |  |  |  |
| SNP-Isoform Pairs | 328,997 | 235,807 | 71.7 | 155,326 | 12,754 | 8.2% |
| Distinct SNPs | 131,804 | 110,154 | 83.6 | 52,186 | 8,746 | 16.8% |
| Distinct isoforms | 1,084 | 826 | 76.2 | 407 | 141 | 34.6% |
| Distinct genes | 590 | 471 | 79.8 | 277 | 98 | 35.4% |

We identify *cis*-sQTLs (at $p < 5 \times 10^{-8}$ ) and *trans*-sQTLs (at $p < 1.5 \times 10^{-13}$ ) in the FHS discovery (n=2,622) and validation (n=1,094) samples ( $p < 1e-4$ ), followed by external replication in 1,020 JHS participants. ‘Eligible pairs’ refer to the number of *cis*- or *trans*-significant pairs in the FHS discovery sample that also meet the inclusion criteria for SNPs and isoforms in JHS.

### External replication of the findings from the FHS using the JHS

Among the 3,588,153 significant *cis-*sQTL-isoform ratio pairs in FHS, 1,583,143 pairs (“eligible pairs”) were available for analysis in JHS, and approximately 23% of these pairs (n=358,654) were replicated (**Table 2**). For significant *trans*-sQTL-isoform ratio pairs in FHS (n=328,997), 155,326 eligible pairs were available in JHS and approximately 8% of these pairs were replicated (**Table 2**). Of the 10,000 most significant *cis-*sQTL-isoform pairs identified in the discovery FHS sample, 88% of the *cis-*pairs and 75% of the *trans-*pairs replicated in the JHS. Of the 100,000 most significant sQTL-isoform pairs identified in the FHS, 69% of the *cis*-pairs and 11% of the *trans*-pairs replicated in the JHS. (**Supplemental Table 4**).

A total of 358,654 *cis*-sQTL-isoform pairs were replicated among eligible pairs in JHS, comprising 2,546 distinct isoforms and 213,050 distinct cis-sQTLs (**Supplemental Table 5**). We identified sentinel cis-sQTLs for each of the 2,546 individual isoforms by defining a sentinel sQTL as the SNP with the lowest p-value for that isoform. We identified 3,048 distinct sentinel cis-sQTLs for these 2,546 isoforms in FHS (**Supplemental Table 5**). We further compared FHS results for these *cis*-sQTLs to those in JHS on the effective allele frequency (EAF) (**Figure 2A**), variance explained (R^2^) in their respective isoform ratios (**Figure 2B**), and effect estimates (**Figure 2C**). We observed a moderate-to-high consistency for EAF (Pearson *r* = 0.69), R^2^ (Pearson *r* = 0.58) and beta (Pearson *r* = 0.74) for these sentinel cis-sQTLs between FHS and JHS (**Figure 2**).

**Figure 2.**
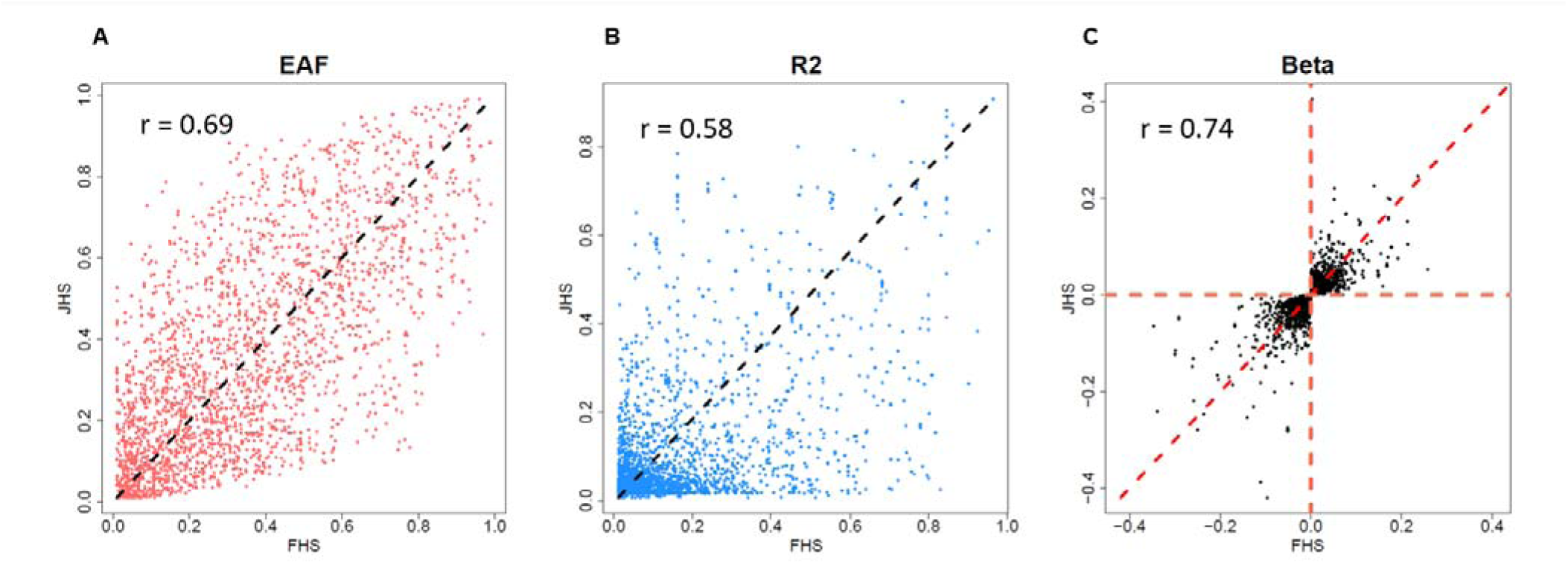
Comparison of effective allele frequency, variance explained and beta estimates of sentinel sQTLs between FHS and JHS. A total of 2,546 distinct isoforms and 213,050 distinct *cis*-sQTLs from the 358,654 common *cis*-sQTL-isoform pairs replicated between FHS and JHS. We identified 3,048 distinct sentinel *cis*-sQTLs for these 2,546 isoforms in FHS. We used Pearson correlation coefficient in each comparison. (**A**) Comparison of effect allele frequency (EAF). (**B**) Comparison of variance of isoform ratio explained (R^2^) by the sentinel cis-sQTLs (**C**). Comparison of effect sizes.

Similarly, we observed differences in allele frequencies and/or effect sizes in the top cis-QTL-isoform pairs replicated between FHS and JHS (**Table 3**). For example, rs10774671 (G>A) in the *OAS1* gene serves as a sentinel *cis*-sQTL for the ENST00000202917.10 transcript in both FHS and JHS. However, EAFs were substantially different between the two cohorts although the beta estimates agree in sign in all but three cases where no value for JHS was available. For the *OAS1* gene, the effect allele frequencies differ between the cohorts (0.62 in FHS versus 0.43 in JHS), and the partial R^2^ (variance explained in the expression ratio of ENST00000202917.10), was 0.95 in FHS versus 0.61 in JHS. Of note, the effect sizes for the association of the isoform ratio of ENST00000202917.10 to *OAS1* were similar in FHS and JHS (−0.26 versus −0.22) (**Table 3**).

**Table 3.** The most significant *cis*-sQTL-isoform pairs identified in the Framingham Heart Study and replication in Jackson Heart Study.

| Sentinel SNP | A:EA | Isoform | Chr | SNP Pos | Gene | Gene Type | FHS |  |  |  | JHS |  |  |  |
| --- | --- | --- | --- | --- | --- | --- | --- | --- | --- | --- | --- | --- | --- | --- |
|  |  |  |  |  |  |  | EAF | R<sup>2</sup> | Beta | -logP | EAF | R<sup>2</sup> | Beta | -logP |
| rs4841 | C:T | ENST00000519690.1 | 5 | 150446963 | *RPS14* | protein_coding | 0.25 | 0.96 | 0.045 | 1859 | 0.2 | 0.91 | 0.014 | 505 |
| rs10774671 | G:A | ENST00000202917.10 | 12 | 112919388 | *OAS1* | protein_coding | 0.62 | 0.95 | -0.258 | 1714 | 0.43 | 0.61 | -0.219 | 203 |
| rs73191241 | A:G | ENST00000388962.4 | 12 | 108619543 | *SELPLG* | protein_coding | 0.19 | 0.92 | 0.095 | 1443 | 0.09 | 0.39 | 0.086 | 108 |
| rs62477626 | T:C | ENST00000475668.6 | 7 | 142065610 | *MGAM* | protein_coding | 0.19 | 0.92 | -0.352 | 1402 | 0.06 | 0.08 | -0.145 | 18 |
| rs7513045 | G:T | ENST00000495526.5 | 1 | 39738494 | *PPIE* | protein_coding | 0.36 | 0.91 | 0.219 | 1314 | 0.19 | 0.85 | 0.21 | 400 |
| rs2318043 | A:G | ENST00000381605.8 | 20 | 1615827 | *SIRPB1* | protein_coding | 0.21 | 0.9 | -0.261 | 1297 | 0.14 | 0.27 | -0.147 | 67 |
| rs41280971 | G:T | ENST00000493091.5 | 7 | 100356834 | *PILRB* | protein_coding | 0.18 | 0.9 | 0.15 | 1284 | 0.14 | 0.06 | 0.038 | 14 |
| rs2425068 | T:C | ENST00000437340.5 | 20 | 35626801 | *CPNE1* | protein_coding | 0.08 | 0.88 | 0.152 | 1169 | 0.01 | 0.51 | 0.079 | 153 |
| rs12898397 | T:C | ENST00000440863.7 | 15 | 74837752 | *ULK3* | protein_coding | 0.6 | 0.87 | -0.263 | 1154 | 0.17 | 0.2 | -0.108 | 50 |
| rs7309653 | A:G | ENST00000541272.1 | 12 | 124910282 | *UBC* | protein_coding | 0.79 | 0.86 | 0.012 | 1103 | 0.27 | 0.85 | 0.055 | 404 |
| rs13320 | C:T | ENST00000368814.8 | 1 | 151874041 | *THEM4* | protein_coding | 0.58 | 0.86 | 0.165 | 1082 | 0.45 | 0.43 | 0.084 | 121 |
| rs14434 | C:T | ENST00000460851.6 | 3 | 150584223 | *EIF2A* | protein_coding | 0.86 | 0.85 | -0.291 | 1045 | 0.62 | 0.88 | -0.049 | 455 |
| rs28374262 | A:G | ENST00000465395.5 | 2 | 218234030 | *ARPC2* | protein_coding | 0.49 | 0.85 | 0.008 | 1044 | 0.08 | 0.41 | 0.027 | 116 |
| rs2724360 | C:T | ENST00000357714.5 | 1 | 207769813 | *CD46* | protein_coding | 0.8 | 0.84 | -0.149 | 1012 | 0.83 | 0.17 | -0.014 | 41 |
| rs2241976 | A:G | ENST00000418251.6 | 2 | 113185893 | *PSD4* | protein_coding | 0.41 | 0.84 | 0.101 | 1011 | 0.69 | 0.2 | 0.029 | 48 |
| rs7063 | A:T | ENST00000296754.7 | 5 | 96774507 | *ERAP1* | protein_coding | 0.3 | 0.83 | 0.165 | 1000 | 0.14 | 0.83 | 0.18 | 375 |
| rs7171507 | T:C | ENST00000564570.1 | 15 | 75444946 | *MAN2C1* | protein_coding | 0.28 | 0.83 | 0.038 | 1000 | 0.44 | 0.58 | 0.038 | 186 |
| rs9616714 | G:A | ENST00000405854.5 | 22 | 49657211 | *C22orf34* | lincRNA | 0.63 | 0.83 | 0.298 | 995 | NA | NA | NA | NA |
| rs452717 | T:G | ENST00000493242.1 | 19 | 54277224 | *LILRB2* | protein_coding | 0.85 | 0.83 | -0.129 | 990 | 0.57 | 0.02 | -0.007 | 6 |
| rs10493821 | C:T | ENST00000489444.6 | 1 | 89009452 | *GBP3* | protein_coding | 0.34 | 0.82 | 0.29 | 963 | 0.08 | 0.54 | 0.226 | 166 |
| rs72494581 | T:C | ENST00000514448.5 | 5 | 181003797 | *BTNL8* | protein_coding | 0.3 | 0.82 | 0.135 | 959 | NA | NA | NA | NA |
| rs372699745 | TA:T | ENST00000343139.9 | 6 | 32593216 | *HLA-DQA1* | protein_coding | 0.29 | 0.82 | -0.208 | 951 | 0.2 | 0.15 | -0.096 | 37 |
| rs7724759 | G:A | ENST00000348386.7 | 5 | 96740783 | *CAST* | protein_coding | 0.33 | 0.82 | 0.1 | 947 | 0.11 | 0.3 | 0.024 | 77 |
| rs11382548 | C:CA | ENST00000334888.9 | 11 | 61398259 | *TMEM216* | protein_coding | 0.85 | 0.81 | 0.147 | 938 | 0.75 | 0.3 | 0.126 | 77 |
| rs34634498 | G:A | ENST00000376228.9 | 6 | 31268623 | *HLA-C* | protein_coding | 0.11 | 0.81 | -0.081 | 924 | 0.1 | 0.28 | -0.083 | 73 |
| rs930232 | G:A | ENST00000263097.9 | 19 | 1036019 | *CNN2* | protein_coding | 0.45 | 0.61 | -0.032 | 532 | 0.41 | 0.026 | -0.027 | 6.44 |
| rs930232 | G:A | ENST00000348419.7 | 19 | 1036019 | *CNN2* | protein_coding | 0.45 | 0.64 | 0.030 | 586 | NA | NA | NA | NA |
| rs5014188 | T:C | ENST00000568865.3 | 19 | 1037109 | *CNN2* | protein_coding | 0.88 | 0.58 | -0.0034 | 481 | 0.76 | 0.24 | -0.014 | 60 |

A:EA, allele:effective allele; EAF, effective allele frequency; R<sup>2</sup>, variance in an isoform ratio that was explained by the sentinel sQTL; -logP, -log10 (p-value)

### Comparison of the set of *cis*-sQTLs and the set of *cis*-eQTLs

We previously reported 6,778,286 significant *cis*-eQTL-eGene pairs from 2,855,111 distinct *cis*-eQTL and 15,982 distinct eGenes (at *p*<5×10^−8^) using the same FHS discovery sample (n=2,622).^23^ Of the sGenes in the present investigation, about 20% (n=974 of 4,971) did not have corresponding eQTL variants (at *p*<5×10^−8^) (**Figure 3A**). We defined the primary sentinel *cis*-sQTL or *cis*-eQTL as the variant with the lowest p-value for each sGene or eGene, respectively. We then compared the explanatory power of the primary sentinel *cis*-sQTL on isoform ratios and the primary sentinel *cis*-eQTL on overall gene expression.

**Figure 3.**
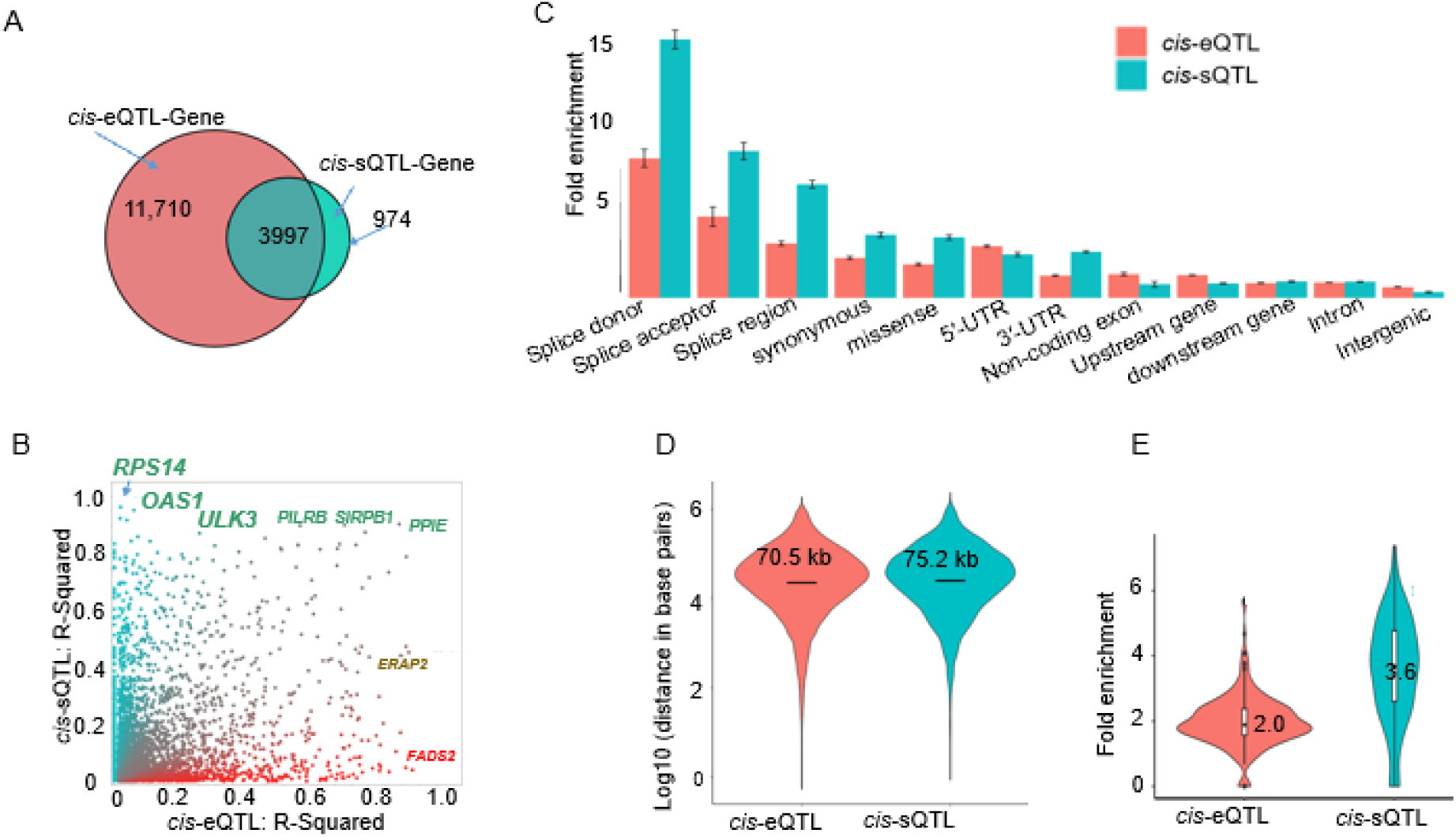
Comparison of cis-sQTLs and cis-eQTLs. (A) Comparison the number of eGenes (targets of cis-eQTLs) and sGenes (targets of cis-sQTLs). (B) Comparison of strength of association, measured as R^2^ (see Methods) for all cis-eQTLs vs. cis-sQTLs. QTLs with nearly exclusive splicing effects are along the Y-axis while those with predominantly expression effects are shown along the X-axis. (C) Comparison of fold enrichment of eQTLs to sQTLs for nominal functional role of SNP, showing the cis-sQTLs are enriched in splice donor/receptor regions compared to cis-eQTLs. (D) Comparison of the location of cis-eQTLs and cis-sQTLs relative to the transcription start site (TSS) of their gene. On average, cis-sQTLs are located 75.2 kb of TSS while cis-eQTLs 70.5 kb of TSS (p<0.05). (E) Comparison of enrichment of eQTLs to that of sQTLs for eCLIP peaks of binding sizes of 203 RNA Binding Proteins from ENCODE.

An isoform is likely to be strongly regulated by genetic variants if its isoform ratio is largely explained (R^2^ >0.8) by its sentinel *cis*-sQTL (**Figure 3B**). Among the 974 sGenes that were not significantly regulated by eQTLs, three had a lead *cis*-sQTL explaining ≥80% of variance in the isoform ratio, 13 had a sentinel *cis*-sQTL explaining 50%-79.9% of variance, and 68 had a sentinel cis-sQTL explaining 20%-49.9% of variance in their respective isoform ratios. For example, rs4841 C>T explained 96% of variance in isoform ENST00000519690.1 of *RPS14*. The rs4841-T allele was associated with a higher ratio of ENST00000519690.1 to overall *RPS14* expression (beta=0.045, -logP =1858.7). We also identified 567,259 SNPs that served as both *cis*-sQTLs and *cis-*eQTLs across 3,384 distinct genes. We compared the variance explained by the sentinel *cis*-sQTLs versus that explained by sentinel *cis*-eQTLs. We found that 85 sentinel SNPs accounted for a substantially higher proportion of variance explained (greater than 50%) as *cis*-sQTLs, than the variance explained as *cis*-eQTLs. This suggests that these sentinel SNPs may have a greater impact on RNA processing than on overall gene expression. For example, rs10774671 G>A in *OAS1* was a strong *cis*-sQTL (R^2^=0.95) for the isoform ratio of ENST00000202917.10, while that same SNP was only a weak eQTL for the overall gene expression level (R^2^=0.05) (**Figure 3B**).

We compared the predicted functions of the sentinel *cis*-eQTLs and *cis*-sQTLs based on their precise physical locations. We found that, compared to *cis*-eQTLs, *cis*-sQTLs were more likely to be enriched in splice donor sites (fold enrichment=14.8 vs. 8.0, logP*>*15), splice acceptor sites (fold change=8.4 vs. 4.7, logP*>*15) (**Figure 3C**). In contrast, eQTLs were more likely than sQTLs to be enriched upstream of a gene (within 5 kb) (fold enrichment=1.31 vs. 0.84, *p* = 0.005) (**Figure 3C**). On average, *cis*-sQTLs are located slightly farther away than *cis*-eQTLs from their respective TSSs (mean distance=75.2 kb vs. 70.5 kb, *p*=0.04) (**Figure 3D**). Additionally, the top *cis*-sQTLs were more enriched in RBP eCLIP peaks than top *cis*-eQTLs (mean fold-enrichment 3.6 vs. 2.0, *p* < 1e-16, see Methods) (**Figure 3E**).

### Enrichment of *cis*-sQTLs in the mQTLs, pQTLs, and GWAS Catalog SNPs

We found that sQTLs were enriched in pQTLs compared to non-sQTL SNPs (fold enrichment=8.0, logP>300) and mQTLs using the GoDMC mQTL database^34^ (fold enrichment=5.7, logP>300).

We also linked *cis*-sQTLs to GWAS Catalog SNPs to investigate whether *cis*-sQTLs are associated with diseases/traits.^33^ *cis*-sQTL variants exhibited enrichment in GWAS Catalog variants for 768 trait categories including blood protein measurement (fold enrichment=7.0, logP>300), BMI-adjusted waist-hip ratio (fold enrichment=6.4, logP>300), and high-density lipoprotein cholesterol measurement (fold enrichment=4.6, logP>300) (**Supplemental Table 6**). In addition, we identified 204 GWAS traits enriched for *trans*-sQTL variants (**Supplemental Table 7**), including BMI-adjusted waist-hip ratio (fold enrichment=16.8, logP>300) and hemoglobin measurement (fold enrichment=10.5, logP>91).

### The relationships between sQTLs, alternative splicing, and diseases/traits

To illustrate the utility of this sQTL resource for identifying putatively causal transcriptome isoforms underlying disease risk, we provide three examples using the top sQTLs identified in our study.

#### The OAS1 gene harbors a strong sentinel sQTL

The *OAS1* (2’-5’-oligoadenylate synthetase 1) gene has long been known to play a role in diabetes and the response to viral infection.^43,44^ The rs10774671 G>A variant, previously identified, ^43,44^ is highly significant in our study, explaining 95% of the variation in the expression ratio of transcript ENST00000202917.10, the second most significant for any gene in our study (**Figure 4A and Table 2**). The role of this sQTL in susceptibility to severe COVID-19 infection was recently described.^45^ *OAS1* contains six exons, encoding seven detectable isoform transcripts in the present study participants (**Supplemental Table 8**), the strongest being ENST00000202917.10 (OAS1-201) and ENST00000452357.6 (*OAS1*-203) (**Figure 4A**). The rs10774671-A allele disrupts the conservative dinucleotide “AG” of the splice acceptor site of intron 5, changing it to “AA” (**Figures 4B & 4C**), with a high probability score of 0.89 for acceptor loss according to SpliceAI.^36^ We investigated RNA-seq data from a randomly selected participant for each of the three genotypes of rs10774671, G/G, A/G, or A/A. The participant with the G/G genotype harbored the canonical “AG” receptor site, resulting in full exclusion of intron 5 in the mature mRNA isoform. Conversely, the participant with the A/A genotype harbored the disrupted receptor site, i.e., from “AG” to “AA,” resulting in retention of intron 5 in mRNA. In contrast, the participant with the A/G genotype had partial retention of intron 5 (**Figure 4C**).

**Figure 4.**
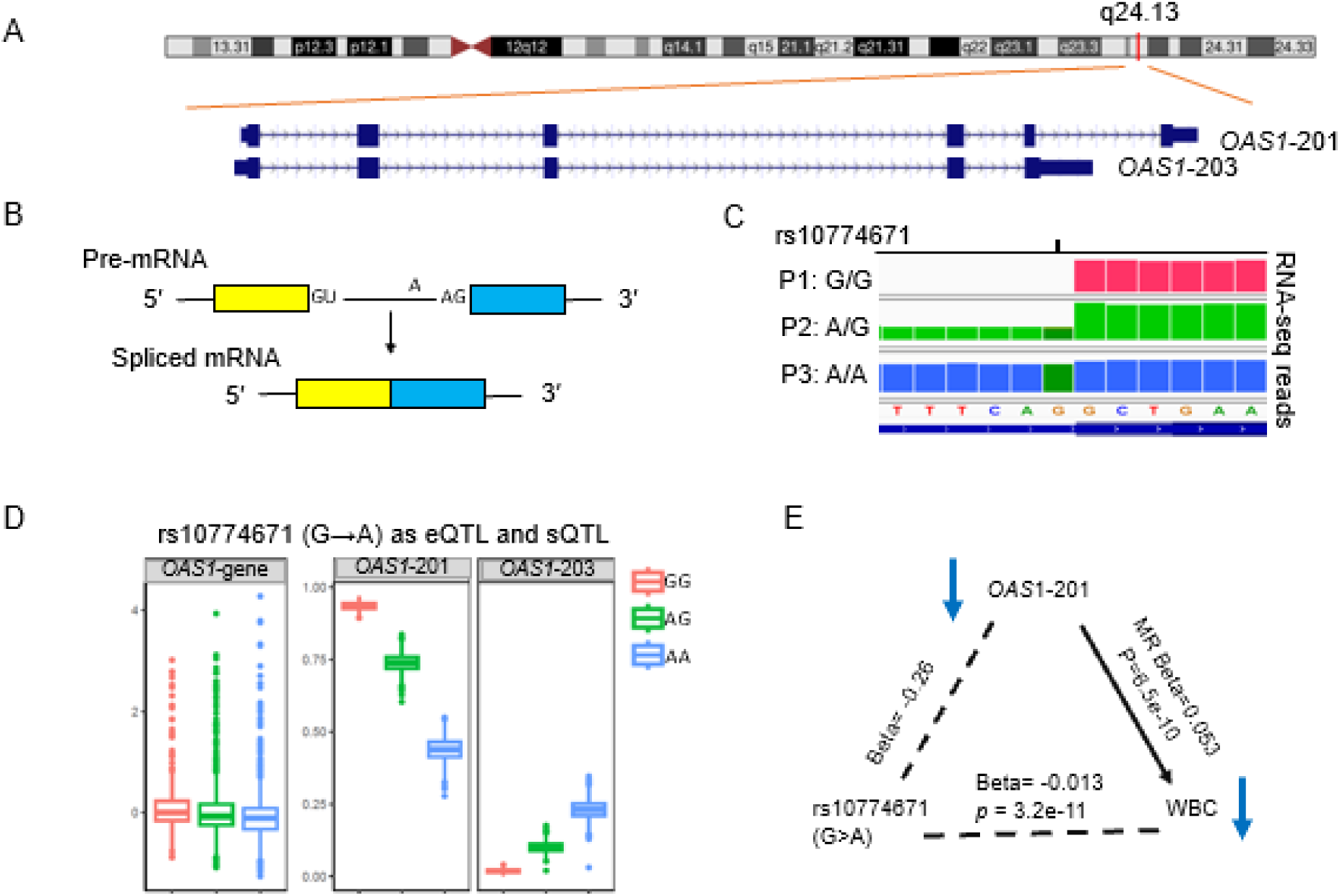
Analyses of sQTLs Illustrated with the *OAS1* Gene. (A). *OAS1* lies on the long arm of chromosome 12 and its expression product (pre-mRNA) is subjected to editing. (B) An intron with a conserved donor sequence ‘GU’ and receptor sequence ‘AG’ is removed from a pre-mRNA, yielding an isoform with the corresponding splice junction. (C) The number of mRNA-seq reads as vertical bars at positions around a potential splice junction mapped to the genome in three FHS participants with rs10774671 (G>A) genotypes G/G, A/G or A/A. These genotypes in the complete absence (G/G), partial absence (A/G), or complete presence (A/A) of splicing, respectively, which in turn result in the high, middle or low ratio levels of the corresponding isoform (*OAS1*-201). (D) Overall expression is weakly affected by rs10774671 genotype while the expression ratio of isoform ENST00000202917.10(*OAS*1-201) and ENST00000452357.6 (*OAS1*-*203*) are strongly affected. (E) rs10774671-A allele of the sentinel *cis*-sQTL is associated with lower levels of ENST00000202917(*OAS*1-201). The lower level of this isoform is causally associated with lower level of white blood cell (WBC) counts by Mendelian randomization and is directionally consistent with the published GWAS finding of rs10774671-A with WBC.

The rs10774671-A variant was strongly associated with a lower isoform ratio of *OAS*1-201 (R^2^=0.95, beta=-0.26, -logP =1713). Conversely, rs10774671-A was associated with higher isoform ratios of the other isoforms including *OAS1*-203 (beta=0.11, -logP=1359) (**Figure 4D**). In addition to its well-known role in modifying COVID-19 severity, the rs10774671-A variant was significantly associated with lower WBC count (beta=-0.013, -logP=10) in a large GWAS^46^ (**Figure 4E**). In MR analysis, a lower isoform ratio of *OAS*1-201(as would be expected with the rs10774671-A allele) was putatively causal for a lower WBC count (MR beta=0.053, -logP=9) (**Supplemental Table 9**).

#### Strong sQTL in the ULK3 gene influence the primary protein structure

*ULK3 (unc-51 like kinase 3)* encodes a serine/threonine protein kinase that serves as a regulator of sonic hedgehog signaling and autophagy.^47,48^ This gene contains 16 exons and encodes 11 detectable isoform transcripts in our study participants (**Supplemental Table 8**). The sentinel *cis*-sQTL, rs12898397 (T>C) is in the 14^th^ exon of *ULK3* (**Table 3**, **Figure 5A**). The ratio of transcript ENST00000440863.7 (*ULK3*-201) was the most significant isoform associated with rs12898397 (T>C).

**Figure 5.**
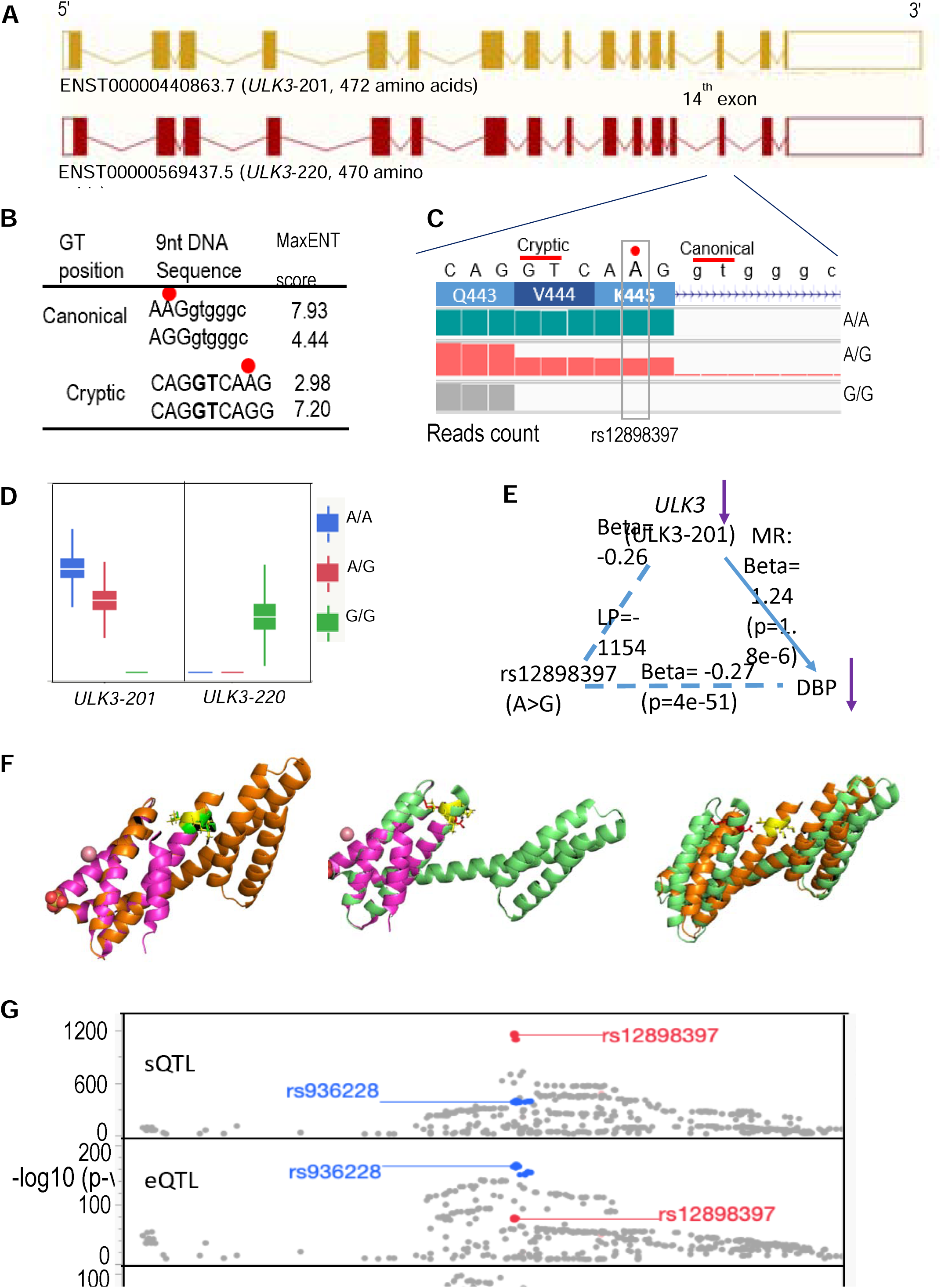
Genetically associated alternative splicing and expression variation in the *ULK3* Gene. **A.** The *ULK3* (unc-51 like kinase 3) on chromosome 15 and resides on the negative DNA strand of the hg38 build of the human genome. It has 16 exons. The isoform ratio of the transcript ENST00000440863.7 (*ULK3*-201) displayed most significant association with the sentinel variant rs12898397 (T>C). This transcript produces a full-length protein with 472 amino acids, whereas isoform ENST00000569437.5 (*ULK3*-220) produces a truncated protein with two amino acids (“VK”) missing at the 5’ of the 14th exon. **B.** The sentinel variant rs12898397 (T>C on the negative strand, A>G on the forward strand) is marked with the red dot. The G allele weakens the canonical 5′GT as a donor (MaxENT score reduced from 7.93 to 4.44) while increasing the chance of using cryptic GT as a donor (score increased from 2.98 to 7.20). **C.** RNA-seq reads are visualized in three FHS subjects (lowercase of nucleotides indicating in an intron region). The rs12898397-T/T genotype (A/A in forward strand) results in the longer isoform ENST00000440863.7, using the canonical 5′GT as donor in splicing. The subject with G/G genotype (on the forward stand) uses the cryptic GT as donor, skipping two codons in the mature isoform. The subject with A/G has a mixed effect: half skipping the two codons. **D.** rs12898397-G is associated with a lower level of isoform ENST00000440863.7 (Table 3). **E.** The lower level of ENST00000440863.7 is causally associated with a lower level of DBP by Mendelian randomization and is directionally consistent with the GWAS association of rs12898397-G with DBP. Likewise, a lower level of this isoform is causally associated with lower Lymphocyte percent, again directionally consistent with the findings within the FHS participants. **F.** Pymol visualization of AlphaFold 3.0-predicted COOH-terminal domains of ULK3 protein variants (resulting from rs12898397 A>G on the forward strand) with and without ‘VK’ is shown. Alignment of the ULK3 crystal structure domain (4WZX) with the 173-aa fragment yields RMSD values of 0.258 (with ‘VK’) and 0.239 with the 171-aa fragment (without ‘VK’). The 173-aa model with ‘VK’ and the 171-aa model without ‘VK’ show conformational differences (RMSD = 5.810) especially in the orientation of the long helix connecting the two 3-helix MIT domains. See Supplemental Materials and Figures 3-4 for details. **G.** The *CYP1A1-ULK3* region is known for its associations with DBP and with lymphocyte percent. This sentinel sQTL is shown in red while the sentinel eQTL is shown in blue. The lower two tracks are the GWAS DBP and lymphocyte percent results from the UKBB data. The sentinel eQTL is slightly stronger for DBP than the sentinel sQTL. Conversely, the sentinel sQTL is stronger for lymphocyte percent than the sentinel eQTL. This suggests that expression and splicing variation may have distinct mechanistic effects on phenotypes.

The *ULK3*-201 produces a full-length protein with 472 amino acids, whereas isoform ENST00000569437.5 (*ULK3*-220) produces a truncated protein with two amino acids (“VK”) missing at the 5’ of the 14^th^ exon (**Figure 5A**). This agrees with the MaxENT prediction, which shows that the rs12898397-C allele greatly weakens the annotated canonical 5’GT donor site (MaxENT score of 7.93 vs. 4.44) while greatly increasing the second GT in the exon as a donor site (MaxENT score of 2.98 vs. 7.20), resulting in the skipping of six nucleotides (“VK” amino acids) due to the rs12898397-C allele (**Figure 5B**). We randomly selected one FHS participants with each of the three genotypes of rs12898397 (T/T, T/C and C/C) and investigated their RNA-seq reads for *ULK3*. As expected, we found that the individual with the rs12898397-T/T genotype used the canonical TG as the donor site to splice out intron 14. In contrast, the individual with the C/C genotype used the cryptic TG site as the donor, resulting in splicing out of a longer intron 14 along with loss of six nucleotides in the mature mRNA, and a corresponding loss of two amino acids in the transcribed protein. The individual with the heterozygous genotype (T/C) displayed half of reads of the six nucleotides compared to that in the individual with T/T genotype (**Figure 5C**). The missing amino acids are located at the C-terminal of the microtubule-Interacting and transport (MIT) domain^49^ (**Supplemental Figure 2**). Google AlphaFold 3 predictions suggested that the absence of the “VK” amino acids in one of the protein isoforms may result in structural differences in the COOH-terminal region (**Figure 4D, Supplemental Figures 3-4**).

The rs12898397-C variant was associated with a lower isoform expression ratio of *ULK3*-201 (R^2^=0.87, beta=-0.26, -logP=1154) (**Table 3**) and a higher isoform expression ratio of *ULK3*-220 (R^2^=0.63, beta=0.16, -logP=573) (**Figure 5D**). We sought to unravel the phenotypic consequences of these two processes (overall gene expression changes versus isoform proportion changes). A higher dosage of the rs12898397-C variant was significantly associated with lower diastolic blood pressure (DBP) in a prior GWAS.^50^ Using rs12898397 as the genetic instrument, we found that a lower expression ratio of *ULK3*-201 level (as would be expected with the rs12898397-C allele) was causally associated with lower DBP (MR beta=1.25, p=1.8e-6) (**Figure 5E**, **Supplemental Table 9**). Similarly, lower expression ratio of *ULK3*-201 was also causally associated with a lower lymphocyte percentage (MR beta=0.054, p=4.9e-11).^46^ Of note, the sentinel eQTL, rs936228, was reported to be associated with comorbidity of COVID-19 critical illness and hypertension.^51^

The *ULK3* gene region harbors several strong *cis*-sQTLs (**Supplemental Table 10**) as well as numerous strong *cis*-eQTLs,^23^ though generally with lower statistical significance compared to the *cis*-sQTLs (**Figure 5F**). We highlighted the SNPs in strong LD (R^2^>0.98) with the sentinel *cis*-sQTL (rs12898397). These SNPs had substantially lower significance as *cis*-eQTLs and were in only moderate LD (R^2^ < 0.63) with the sentinel *cis*-eQTL. Similarly, SNPs in strong LD (R^2^>0.9) with the sentinel *cis*-eQTL had lower significance as *cis*-sQTLs and were in only moderate LD (R^2^ <0.63) with the sentinel *cis*-sQTL, despite all being located within the *ULK3* gene region.

#### Different sQTLs are associated with multiple isoform transcripts of CNN2

*CNN2* (Calponin 2, 19p13.3) encodes a regulatory protein that interacts through its three well-conserved 26 amino acid residue domains, with actin, tropomyosin and other cellular components to modify cytoskeleton dynamics.^52,53^. *CNN2* consists of five exons, producing seven detectable isoform transcripts in our study participants (**Figure 6A**). We found two significant sentinel *cis*-sQTLs (**Supplemental Table 2**), rs930232 (G>A) and rs5014188 (T>C) (pairwise LD, R^2^=0.13, all population in LDlink^54^), that were linked to three *CNN2* isoforms, ENST00000263097.9 (*CNN2*-201), ENST00000348419.7 (*CNN2*-202), and ENST00000568865.3 (*CNN2*-209) (**Figure 6B**).

**Figure 6.**
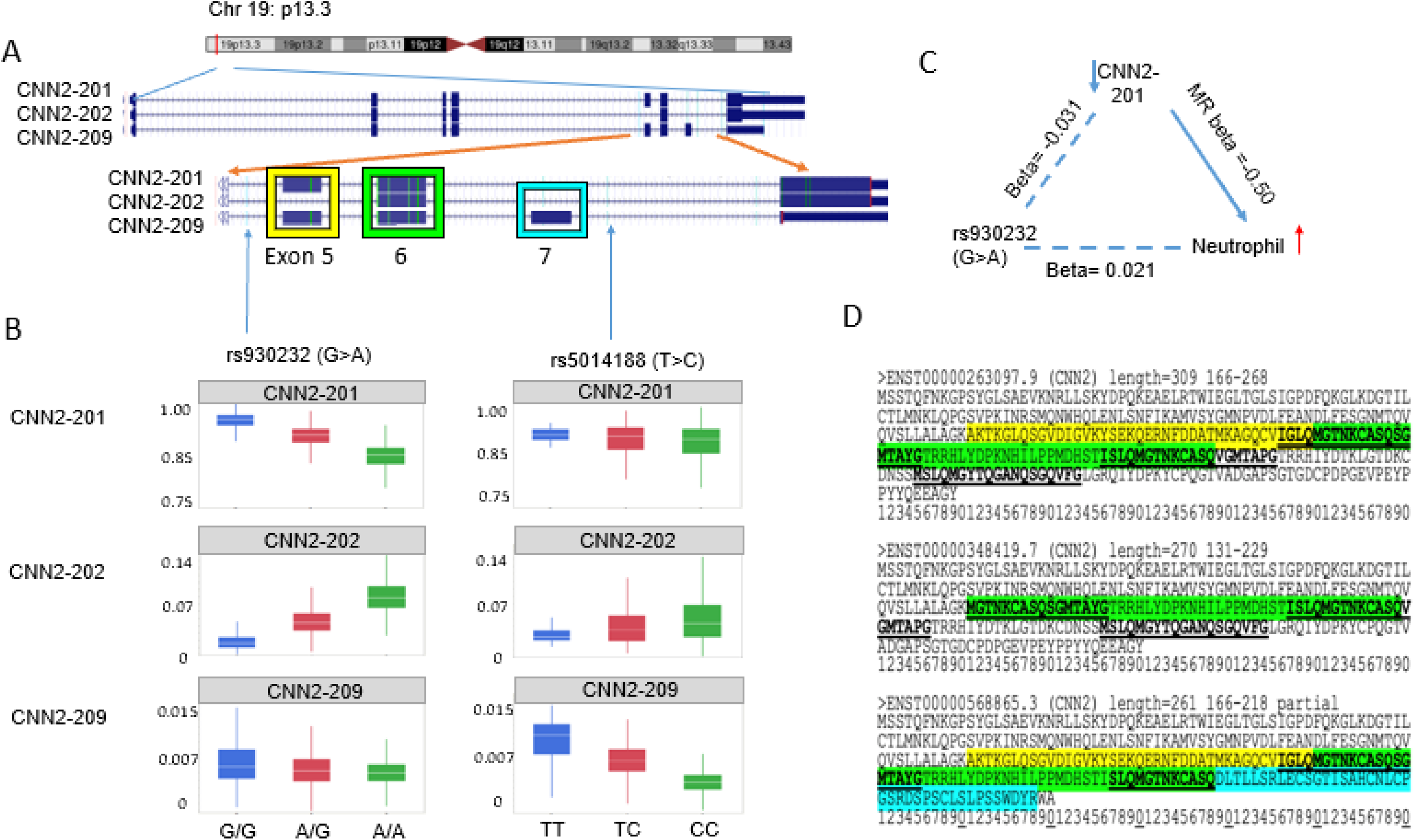
*CNN2* transcripts under genetic control. Three transcript expression ratios, ENST00000263097.9 (*CNN2*-201), ENST00000348419.7 (*CNN2*-202), and ENST00000568865.3 (*CNN2*-209) are under significant genetic control and differ by inclusion of Exons 5 and 6, Exon 6 alone, or Exons 5, 6 and 7, resp. **A)** Genome view of *CNN2* on Chr 19. **B)** The expression ratios are strongly associated with genotype at the sentinel sQTL rs930232 for *CNN2*-201 and *CNN2*-202. The expression ratio of *CNN2*-209 is more strongly associated with the sentinel sQTL, rs5014188 than are the other two expression ratios. **C)** rs930232 (G>A) lowers expression of *CNN2*-201 which in turn raises neutrophil percent (proven by Mendelian randomization), which aligns with the observed positive association of rs930232 with neutrophil percent. **D)** Predicted protein sequence of calponin 2 isoforms highlighting the contributions from Exons 5, 6 and 7. The three conserved 26 residue domains involved in binding of calponin to actin are underlined in each isoform.

The rs930232-A allele was identified as a strong *cis*-sQTL, associated with a lower isoform ratio of *CNN2*-201 (R^2^=0.61, beta=-0.031, -logP=531) and with higher ratio of *CNN2*-202 (R2=0.65, beta=0.030, -logP=586); however, it was only weakly associated with *CNN2*-209 (R^2^=0.04, beta=-0.00064, -logP=27) (**Supplemental Table 2**). Despite being a strong sQTL, the rs930232 (G>A) variant was not predicted to directly influence splice sites by either SpliceAI or MaxENT. The rs930232-A allele was associated with a higher neutrophil count (beta=0.021, -logP=26) in 746,667 individuals in a blood-cell composition GWAS (**Figure 6C**).^55^ In FHS participants, the *CNN2*-201 isoform ratio was negatively associated with neutrophil count (beta=-0.00042, *p*=0.0015); MR also found that *CNN2*-201 isoform ratio was negatively associated with neutrophil count (beta=-0.50, -logP=14.7) (**Figure 6C**). rs5014188-C was strongly associated with a lower relative level of *CNN2*-209 (R^2^=0.58, beta=-0.0034, -logP=482) while it showed no significant association with CNN2-201 (*p*>5e-8) and a much weaker, positive association with CNN2-202 (R^2^=0.02, beta=0.0081, -logP= 13) (**Figure 6B**). The effect of *CNN2* isoform variation on predicted protein sequence is depicted in **Figure 6D**. Predicted protein sequences were based on the standard translation of each of the mRNA sequences in the UCSC Genome Browser database^56^ (https://genome.ucsc.edu/index.html). Three well-known calponin-like repeat motifs are indicated, superimposed on the contributions from exons 5, 6, and 7 in the three protein isoforms.^52^ Loss of exon 5 in *CNN2*-202 truncates the first repeat, while the gain of exon 7 in *CNN2*-209 truncates the second repeat and obliterates the third repeat entirely.^52^ These motifs are essential to the actin-binding properties of calmodulin 2, and thus, modifications of the relative amounts of these three isoforms, *CNN2*-201, *CNN2*-202, and *CNN2*-209, may mediate observed phenotypic consequences of *CNN2* sQTLs.

## DISCUSSION

We conducted comprehensive sQTL analyses using whole-genome and whole-transcriptome sequencing to generate a catalog of genetic variants associated with isoform variation genome wide using whole blood DNA and RNA from a large study sample. Using the discovery set of 2622 FHS participants, we identified 3,588,153 significant *cis*-sQTL-isoform pairs (p<5e-8), with a validation rate of 61% (p< 1e-4) in the FHS validation set (n=1,094). External replication in 1,020 JHS participants revealed replication rates of 86% for the 10,000 most significant sQTL-isoform ratio pairs (comprising 155 sGenes) and 69% for the 100,000 most significant pairs (comprising 655 sGenes) identified in FHS discovery. We distinguished *cis*-sQTLs that did not show substantial impact on overall gene expression levels from those that were also *cis*-eQTLs by comparing several key properties. To illustrate the impact of sQTLs on splice variation, we explored in detail the properties of one sQTL that disrupts a canonical splice site, and another that introduces a cryptic splice site, leading to AS. By integrating GWAS Catalog data with sQTLs,^33^ we observed an enrichment of GWAS SNPs within sQTLs. Furthermore, we found that several top isoform transcripts were causally associated with CVD traits or WBC composition, providing insights into the possible influence of genetic variations on disease susceptibility through regulation of RNA splicing. Our findings advance understanding of the genetic control of RNA splicing, underscoring the importance of sQTL mapping for elucidating SNP-trait associations.

Transcription isoforms can arise through diverse molecular processes such as alternative splicing, alternative cleavage and polyadenylation, alternative promoter usage, and post-translational modifications. To provide proof-of-concept of the contributions of *cis-*sQTLs to AS, we annotated the sentinel *cis*-sQTLs using SpliceAI^36^ and MaxEnt ^37^ methods. These approaches use different algorithms and predict different aspects of SNP effects on AS.^36,57^ SpliceAI predicts that rs107746 (G>A) of the *OAS1* gene (Figure 3) increases the probability of 3’ splice acceptor loss by 89%, while the delta MaxENT score for this site is low. In contrast, MaxENT predicts that the *ULK3* rs12898397 (T>C) variant disrupts the annotated canonical 5’GT as a donor while greatly strengthening the second GT in the exon as a donor. The effects predicted by the two methods can be validated using RNA-seq data. Leveraging RNA-seq data, we visualized the effects of rs107746 (G>A) and rs12898397 (T>C) on transcript splicing by demonstrating the distinct RNA-seq read patterns in randomly selected FHS participants with the three genotypes of each SNP. Although leveraging RNA-seq data to investigate the direct influence of *cis*-sQTLs will help understand the direct impact of an *cis*-sQTL in relation to RNA splicing, a systematic approach^58,59^ should be applied to characterize the effects of the significant *cis*-sQTLs on RNA splicing across the human genome.

We found that *cis*-sQTLs are enriched in GWAS SNPs associated with complex traits, which is consistent with previous studies.^13,14,17^ Linking to SNPs in the GWAS Catalog,^33^ we found cis-sQTL variants are enriched in 768 trait categories, with significant fold enrichment for major cardiovascular risk factors like BMI-adjusted waist-hip ratio (6.4-fold, -logP>300) and HDL cholesterol (4.6-fold, -LogP>300), consistent to what was previously reported.^23^ Of note, only 0.6% of GWAS Catalog SNPs were annotated as splicing related, suggesting that many GWAS SNPs with unknown functions might directly affect RNA splicing, and that splice effects may underlie the observed SNP-trait associations. As proof of concept, we conducted MR to infer causal associations between isoform transcript levels and CVD risk factors and WBC composition. For example, the sentinel *cis*-sQTL rs10774671 in *OAS1* has been associated with COVID-19 severity in large GWAS,^60,61^ and implicated in the COVID-19 causal pathway, with rs10774671-G providing protection.^62–64^ Along with *OAS2* and *OAS3*, *OAS1* encodes one of the three antiviral 2′,5′-oligoadenylate synthetase (OAS) enzymes, which are interferon-inducible antiviral proteins activating the latent form of ribonuclease L (RNase L).^65^ rs10774671-G leads to prenylation of the protein isoform, *OAS1*-p46. This prenylated protein isoform targets the endomembrane system, enhancing the binding of *OAS1*-p46 to the double-stranded viral RNA and initiates RNA degradation of the viral RNA through an RNAase L pathway.^45,66^ Disruption of the 3’ receptor site by rs10774671-A leads to the retention of the fifth intron in RNA splicing and the production of *OAS1*-p42. Individuals with the *OAS1*-p42 protein isoform are more susceptible to viral infection.^45,66^ The rs10774671 (G>A) variant is pleotropic. It was also linked to type I diabetes, with the G allele increasing the odds of the condition.^67^ In large GWAS studies of WBC composition, rs10774671-A was additionally associated with a lower WBC count.^55^ Through MR analysis, we found that the rs10774671-A variant is causally linked to a lower WBC count via regulation of AS, which is consistent with the association of rs10774671-A with WBC count in GWAS (**Figure 4**).

We provided an additional example to illustrate the potential of sQTLs to regulate splice variation. In the *ULK3* gene, the sentinel sQTL rs12898397 (T>C) is associated with two phenotypes in GWAS, blood pressure^68^ and lymphocyte proportion.^46^ The *ULK3*-coded unc-51 like kinase 3 is a serine/threonine protein kinase that is widely expressed in multiple tissues, especially tissues that require high energy, including brain, liver, and kidney.^47,48^ The rs12898397-C variant disrupts the canonical donor splice site in *ULK3*, shifting the splice site from chr15:74,837,750 to a competing site at chr15:74,837,755 (Figure 4). This leads to the skipping of 6 bp (corresponding to amino acids VK” in the mature mRNA, producing the truncated isoform ENST00000569437.5 (ULK3-220), which encodes a protein with 470 amino acids (**Figure 5**). The absence of “VK” at the C-terminal of the MIT domain,^49^ which is involved in organelles and vesicles trafficking, disrupts ESCRT (endosomal sorting complexes required for transport) – mediated separation of daughter cells during cytokinetic abscission.^69–71^ Using Google AlphaFold 3.0^38,39^ predictions, we suggest that the truncated ULK3 variant has a different structure compared to the full-length variant (**Figure 5F**) perhaps relevant to its function mediated through binding of its MIT domain to ESCRT-III MIM subunits.^71^ Moreover, many SNPs in the same locus are significant sQTLs for *ULK3* (**Figure 5**). The sQTLs for *ULK3* explain a substantial proportion of variance in their respective isoform transcript ratios; in contrast, they are much weaker eQTLs for overall *ULK3* expression. Additional research is needed to compare multiple sQTLs and eQTLs underlying observed trait associations.

We illustrated the additional complexity that can be revealed through the example of the *CNN2* gene, which along with the homologous genes *CNN1* and *CNN3,* encodes calponin isoforms 1, 2, and 3, respectively, executing different physiological roles based on their cell type-specific expression.^72–74^ The protein products of the 12 observed *CNN2* isoform transcripts interact with several cellular components, including actin and tropomyosin.^52^ Calponin proteins are implicated in the structural arrangement of actin filaments and may contribute to smooth muscle contraction and cell adhesion.^52^ In the present study, the rs930232 G>A variant was significantly associated with two major isoform transcripts, ENST00000263097.9 (*CNN2*-201) and ENST00000348419.7 (*CNN2*-202), with opposite directionality. These two transcripts differ by the inclusion of exon 5, in *CNN2*-201 its exclusion in *CNN2*-202 (http://useast.ensembl.org) (**Figure 6**). The calponin repeat motifs, of which there are three in the native protein, are the modules involved in binding of this protein to actin and other proteins.^52^ These modules are partially disrupted by the loss of exon 5, which likely alters the functionality of *CNN2* protein. A third transcript variant, ENST00000568865.3 (*CNN2*-209), includes an additional exon and is strongly associated with rs5014188 (T>C). The effect of inclusion of this exon is to further disrupt the repeat motifs, and potentially impair protein function.^52^ Alterations in the relative levels of the first two transcript variants are associated with neutrophil percent and neutrophil count (**Figure 6**). While there are eQTLs for *CNN2* that are in moderate LD with the lead sQTL, it is reasonable to infer that splicing variation plays a role in neutrophil effects of this gene.

We identified approximately 3.5 million cis-sQTL-isoform pairs in our discovery sample. Whereas the overall internal validation rate was 61%, the external replication rate in JHS was only 23%. In contrast, the external validation rate in JHS was 88% and 69% for the top 10,000 and 100,000 *cis*-sQTL-transcript ratio pairs from discovery. This discrepancy likely arises from differences in sample size and RNA extraction sources. The FHS sample used for internal validation was slightly over half the size of the discovery sample, while the JHS sample size was less than half, which may partially explain the lower replication rate. Additionally, RNA was extracted from whole blood in the FHS, leading to a more complex RNA profile due to the presence of all blood cell types, including all white blood cells and platelets. In contrast, the JHS used PBMCs, provided a more targeted RNA profile to immune cells (lymphocytes and monocytes).^75^ Moreover, genetic background also plays a role, as FHS participants were predominantly of European ancestry (median EUR similarity was >90%), while JHS participants were of African ancestry (median AFR genetic similarity >80%); in FHS median EUR similarity was >90%.^25^ Replication of our results in additional study samples is warranted.

We compared sQTLs/sGenes with eQTLs/eGenes from our previous study with the same FHS participants.^23^ We found that SNPs acting as both eQTLs and sQTLs affect overall gene expression and isoform-to-transcript ratios differently, suggesting distinct impacts or different causal variants.^17^ Approximately 80% of sGenes are also eGenes, representing a higher sGene-to-eGene overlap ratio compared to previous studies,^13,14,17^ likely due to our larger sample size (n=2,622) and different statistical methods for identifying sQTLs. Unlike methods using junction read counts,^76,77^ our isoform-ratio method measures the ratio of isoform to total gene expression,^15^ which may better capture functional changes but could be less accurate for unknown isoforms. Additionally, our stringent significance threshold (p<5e-8) in eQTL^23^ and sQTL identification differs from the more liberal FDR approach^13,14^ used in most other studies, which could affect overlap ratios. Additionally, applying a Hardy-Weinberg equilibrium (HWE) filter in admixed cohorts like JHS can be problematic, as many ancestry-differentiated SNPs may not meet HWE criteria in these populations. The investigator only considered “common” genetic variants (MAF > 0.01), so further investigation into “secondary” splicing QTLs is warranted.

In summary, our comprehensive investigation sheds light on the role of genetic variation in AS and isoform variation. These findings significantly enhance our understanding of genetic regulation of RNA splicing, highlighting the significant role of sQTL mapping in unraveling SNP-trait associations.

## Supporting information

Supplemental Methods and Results

## Data availability and materials

The FHS datasets analyzed in the present study are available at the dbGAP repository phs000007.v32.p13 (https://www.ncbi.nlm.nih.gov/projects/gap/cgi-bin/study.cgi?study_id=phs000007.v30.p11). For JHS, datasets are available from coordinating center by request or through dbGaP accessions phs000964/phs002256.

## Reference

1. Pan, Q., Shai, O., Lee, L.J., Frey, B.J. & Blencowe, B.J. Deep surveying of alternative splicing complexity in the human transcriptome by high-throughput sequencing. Nat Genet 40, 1413–5 (2008).

2. Chow, L.T., Gelinas, R.E., Broker, T.R. & Roberts, R.J. An amazing sequence arrangement at the 5’ ends of adenovirus 2 messenger RNA. Cell 12, 1–8 (1977).

3. Scotti, M.M. & Swanson, M.S. RNA mis-splicing in disease. Nat Rev Genet 17, 19–32 (2016).

4. Wang, G.S. & Cooper, T.A. Splicing in disease: disruption of the splicing code and the decoding machinery. Nat Rev Genet 8, 749–61 (2007).

5. Kornblihtt, A.R. et al. Alternative splicing: a pivotal step between eukaryotic transcription and translation. Nat Rev Mol Cell Biol 14, 153–65 (2013).

6. Shayevitch, R., Askayo, D., Keydar, I. & Ast, G. The importance of DNA methylation of exons on alternative splicing. RNA 24, 1351–1362 (2018).

7. Barash, Y. et al. Deciphering the splicing code. Nature 465, 53–9 (2010).

8. Battle, A. et al. Characterizing the genetic basis of transcriptome diversity through RNA-sequencing of 922 individuals. Genome Res 24, 14–24 (2014).

9. Lappalainen, T. et al. Transcriptome and genome sequencing uncovers functional variation in humans. Nature 501, 506–11 (2013).

10. Takata, A., Matsumoto, N. & Kato, T. Genome-wide identification of splicing QTLs in the human brain and their enrichment among schizophrenia-associated loci. Nat Commun 8, 14519 (2017).

11. Raj, T. et al. Integrative transcriptome analyses of the aging brain implicate altered splicing in Alzheimer’s disease susceptibility. Nat Genet 50, 1584–1592 (2018).

12. Tian, J. et al. CancerSplicingQTL: a database for genome-wide identification of splicing QTLs in human cancer. Nucleic Acids Res 47, D909–D916 (2019).

13. Consortium, G.T. The GTEx Consortium atlas of genetic regulatory effects across human tissues. Science 369, 1318–1330 (2020).

14. Garrido-Martin, D., Borsari, B., Calvo, M., Reverter, F. & Guigo, R. Identification and analysis of splicing quantitative trait loci across multiple tissues in the human genome. Nat Commun 12, 727 (2021).

15. Yamaguchi, K. et al. Splicing QTL analysis focusing on coding sequences reveals mechanisms for disease susceptibility loci. Nat Commun 13, 4659 (2022).

16. He, Y., Huang, L., Tang, Y., Yang, Z. & Han, Z. Genome-wide Identification and Analysis of Splicing QTLs in Multiple Sclerosis by RNA-Seq Data. Front Genet 12, 769804 (2021).

17. Qi, T. et al. Genetic control of RNA splicing and its distinct role in complex trait variation. Nat Genet 54, 1355–1363 (2022).

18. GTEx Consortium. The GTEx Consortium atlas of genetic regulatory effects across human tissues. Science 369, 1318–1330 (2020).

19. Wen, J. et al. Gene expression and splicing QTL analysis of blood cells in African American participants from the Jackson Heart Study. Genetics 228(2024).

20. Splansky, G.L. et al. The Third Generation Cohort of the National Heart, Lung, and Blood Institute’s Framingham Heart Study: design, recruitment, and initial examination. Am J Epidemiol 165, 1328–35 (2007).

21. Kannel, W.B., Feinleib, M., McNamara, P.M., Garrison, R.J. & Castelli, W.P. An investigation of coronary heart disease in families. The Framingham offspring study. Am J Epidemiol 110, 281–90 (1979).

22. Wilson, J.G. et al. Study design for genetic analysis in the Jackson Heart Study. Ethn Dis 15, S6–30-37 (2005).

23. Liu, C. et al. Whole genome DNA and RNA sequencing of whole blood elucidates the genetic architecture of gene expression underlying a wide range of diseases. Sci Rep 12, 20167 (2022).

24. Joehanes, R. et al. Integrated genome-wide analysis of expression quantitative trait loci aids interpretation of genomic association studies. Genome Biol 18, 16 (2017).

25. Taliun, D. et al. Sequencing of 53,831 diverse genomes from the NHLBI TOPMed Program. Nature 590, 290–299 (2021).

26. Lun, A.T., Chen, Y. & Smyth, G.K. It’s DE-licious: A Recipe for Differential Expression Analyses of RNA-seq Experiments Using Quasi-Likelihood Methods in edgeR. Methods Mol Biol 1418, 391–416 (2016).

27. Robinson, M.D. & Oshlack, A. A scaling normalization method for differential expression analysis of RNA-seq data. Genome Biol 11, R25 (2010).

28. Chen, Y., Lun, A.T. & Smyth, G.K. From reads to genes to pathways: differential expression analysis of RNA-Seq experiments using Rsubread and the edgeR quasi-likelihood pipeline. F1000Res 5, 1438 (2016).

29. Liu, C., et al. Whole Genome DNA and RNA Sequencing of Whole Blood Elucidates the Genetic Architecture of Gene Expression Underlying a Wide Range of Diseases. medRxiv (2022).

30. Wen, J. et al. Gene Expression and Splicing QTL Analysis of Blood Cells in African American Participants from the Jackson Heart Study. bioRxiv (2023).

31. Consortium, E.P. An integrated encyclopedia of DNA elements in the human genome. Nature 489, 57–74 (2012).

32. Luo, Y. et al. New developments on the Encyclopedia of DNA Elements (ENCODE) data portal. Nucleic Acids Res 48, D882–D889 (2020).

33. Sollis, E. et al. The NHGRI-EBI GWAS Catalog: knowledgebase and deposition resource. Nucleic Acids Res 51, D977–D985 (2023).

34. Min, J.L. et al. Genomic and phenotypic insights from an atlas of genetic effects on DNA methylation. Nat Genet 53, 1311–1321 (2021).

35. Sun, B.B. et al. Plasma proteomic associations with genetics and health in the UK Biobank. Nature 622, 329–338 (2023).

36. Jaganathan, K. et al. Predicting Splicing from Primary Sequence with Deep Learning. Cell 176, 535–548 e24 (2019).

37. Yeo, G. & Burge, C.B. Maximum entropy modeling of short sequence motifs with applications to RNA splicing signals. J Comput Biol 11, 377–94 (2004).

38. Abramson, J. et al. Accurate structure prediction of biomolecular interactions with AlphaFold 3. Nature 630, 493–500 (2024).

39. Jumper, J. et al. Highly accurate protein structure prediction with AlphaFold. Nature 596, 583–589 (2021).

40. Berman, H.M. et al. The Protein Data Bank. Nucleic Acids Res 28, 235–42 (2000).

41. Magala, P., Klevit, R.E., Thomas, W.E., Sokurenko, E.V. & Stenkamp, R.E. RMSD analysis of structures of the bacterial protein FimH identifies five conformations of its lectin domain. Proteins 88, 593–603 (2020).

42. Hemani, G. et al. The MR-Base platform supports systematic causal inference across the human phenome. Elife 7(2018).

43. Field, L.L. et al. OAS1 splice site polymorphism controlling antiviral enzyme activity influences susceptibility to type 1 diabetes. Diabetes 54, 1588–91 (2005).

44. Bonnevie-Nielsen, V. et al. Variation in antiviral 2’,5’-oligoadenylate synthetase (2’5’AS) enzyme activity is controlled by a single-nucleotide polymorphism at a splice-acceptor site in the OAS1 gene. Am J Hum Genet 76, 623–33 (2005).

45. Wickenhagen, A. et al. A prenylated dsRNA sensor protects against severe COVID-19. Science 374, eabj3624 (2021).

46. Vuckovic, D. et al. The Polygenic and Monogenic Basis of Blood Traits and Diseases. Cell 182, 1214–1231 e11 (2020).

47. Rubin, L.L. & de Sauvage, F.J. Targeting the Hedgehog pathway in cancer. Nat Rev Drug Discov 5, 1026–33 (2006).

48. Luo, S., Zheng, N. & Lang, B. ULK4 in Neurodevelopmental and Neuropsychiatric Disorders. Front Cell Dev Biol 10, 873706 (2022).

49. Ciccarelli, F.D. et al. The identification of a conserved domain in both spartin and spastin, mutated in hereditary spastic paraplegia. Genomics 81, 437–41 (2003).

50. Keaton, J.M. et al. Genome-wide analysis in over 1 million individuals of European ancestry yields improved polygenic risk scores for blood pressure traits. Nat Genet 56, 778–791 (2024).

51. Guo, H., Li, T. & Wen, H. Identifying shared genetic loci between coronavirus disease 2019 and cardiovascular diseases based on cross-trait meta-analysis. Front Microbiol 13, 993933 (2022).

52. Liu, R. & Jin, J.P. Calponin isoforms CNN1, CNN2 and CNN3: Regulators for actin cytoskeleton functions in smooth muscle and non-muscle cells. Gene 585, 143–153 (2016).

53. Plagnol, V., Smyth, D.J., Todd, J.A. & Clayton, D.G. Statistical independence of the colocalized association signals for type 1 diabetes and RPS26 gene expression on chromosome 12q13. Biostatistics 10, 327–34 (2009).

54. Machiela, M.J. & Chanock, S.J. LDlink: a web-based application for exploring population-specific haplotype structure and linking correlated alleles of possible functional variants. Bioinformatics 31, 3555–7 (2015).

55. Chen, M.H. et al. Trans-ethnic and Ancestry-Specific Blood-Cell Genetics in 746,667 Individuals from 5 Global Populations. Cell 182, 1198–1213 e14 (2020).

56. Nassar, L.R. et al. The UCSC Genome Browser database: 2023 update. Nucleic Acids Res 51, D1188–D1195 (2023).

57. Bailey, T.L. & Elkan, C. Fitting a mixture model by expectation maximization to discover motifs in biopolymers. Proc Int Conf Intell Syst Mol Biol 2, 28–36 (1994).

58. Liu, L., Das, U., Ogunsola, S. & Xie, J. Transcriptome-Wide Detection of Intron/Exon Definition in the Endogenous Pre-mRNA Transcripts of Mammalian Cells and Its Regulation by Depolarization. Int J Mol Sci 23(2022).

59. Khodor, Y.L., Menet, J.S., Tolan, M. & Rosbash, M. Cotranscriptional splicing efficiency differs dramatically between Drosophila and mouse. RNA 18, 2174–86 (2012).

60. Zhou, S. et al. A Neanderthal OAS1 isoform protects individuals of European ancestry against COVID-19 susceptibility and severity. Nat Med 27, 659–667 (2021).

61. Huffman, J.E. et al. Multi-ancestry fine mapping implicates OAS1 splicing in risk of severe COVID-19. Nat Genet 54, 125–127 (2022).

62. Banday, A.R. et al. Genetic regulation of OAS1 nonsense-mediated decay underlies association with COVID-19 hospitalization in patients of European and African ancestries. Nat Genet 54, 1103–1116 (2022).

63. Pairo-Castineira, E. et al. Genetic mechanisms of critical illness in COVID-19. Nature 591, 92–98 (2021).

64. Initiative, C.-H.G. Mapping the human genetic architecture of COVID-19. Nature 600, 472–477 (2021).

65. Sadler, A.J. & Williams, B.R. Interferon-inducible antiviral effectors. Nat Rev Immunol 8, 559–68 (2008).

66. Soveg, F.W. et al. Endomembrane targeting of human OAS1 p46 augments antiviral activity. Elife 10(2021).

67. Tessier, M.C. et al. Type 1 diabetes and the OAS gene cluster: association with splicing polymorphism or haplotype? J Med Genet 43, 129–32 (2006).

68. Evangelou, E. et al. Genetic analysis of over 1 million people identifies 535 new loci associated with blood pressure traits. Nat Genet 50, 1412–1425 (2018).

69. Wenzel, D.M. et al. Comprehensive analysis of the human ESCRT-III-MIT domain interactome reveals new cofactors for cytokinetic abscission. Elife 11(2022).

70. Iwaya, N. et al. A common substrate recognition mode conserved between katanin p60 and VPS4 governs microtubule severing and membrane skeleton reorganization. J Biol Chem 285, 16822–9 (2010).

71. Caballe, A. et al. ULK3 regulates cytokinetic abscission by phosphorylating ESCRT-III proteins. Elife 4, e06547 (2015).

72. Gao, J., Hwang, J.M. & Jin, J.P. Complete nucleotide sequence, structural organization, and an alternatively spliced exon of mouse h1-calponin gene. Biochem Biophys Res Commun 218, 292–7 (1996).

73. Strasser, P., Gimona, M., Moessler, H., Herzog, M. & Small, J.V. Mammalian calponin. Identification and expression of genetic variants. FEBS Lett 330, 13–8 (1993).

74. Applegate, D., Feng, W., Green, R.S. & Taubman, M.B. Cloning and expression of a novel acidic calponin isoform from rat aortic vascular smooth muscle. J Biol Chem 269, 10683–90 (1994).

75. Joehanes, R. et al. Gene expression analysis of whole blood, peripheral blood mononuclear cells, and lymphoblastoid cell lines from the Framingham Heart Study. Physiol Genomics 44, 59–75 (2012).

76. Li, Y.I. et al. Annotation-free quantification of RNA splicing using LeafCutter. Nat Genet 50, 151–158 (2018).

77. Pervouchine, D.D., Knowles, D.G. & Guigo, R. Intron-centric estimation of alternative splicing from RNA-seq data. Bioinformatics 29, 273–4 (2013).

