## Supplemental Methods and Results for "Integrating Whole Genome and Transcriptome Sequencing to Characterize the Genetic Architecture of Isoform Variation and its Implications for Health and Disease"

**Limited external replication in GTEx**

Although GTEx has produced an extensive resource for sQTL analysis,(13, 14) we conducted limited replication using the GTEx data owning to significant methodological differences between the FHS and GTEx approaches in identifying sQTLs. We compared our sQTL results with those generated by Garrido-Martin et al. using 670 samples with GTEx WGS and RNA-seq data,(14) focusing on sQTL-sGene pairs due to methodological differences (14) (see Methods). Of the 1,994,197 sQTL-sGene pairs in our study, 9% were replicated in the Garrido-Martin resource(14) (**Supplemental Table 11**). The replication rate increased significantly with stricter criteria. Of the 1,006,396 sQTL-sGene pairs with MAF>0.2, 45% replicated within the Garrido-Martin results. Among the top 10% of pairs in our study with MAF >0.2 and -log10 (p) >108 (corresponding to R^2^ >18%), 70% replicated among the 135,882 pairs in the Garrido-Martin resource (**Supplemental Table 11**).

**Prediction of protein structures by Google AlphaFold 3.0**

We used Google AlphaFold 3.0 to predict protein structures^38,39^ for two protein coding transcript isoforms associated with the sQTL variant rs12898397 [T>C] in *ULK3* [unc-51 like kinase 3]) as an example of how such variants might affect protein structures (**Supplemental Materials**). The sQTL variant and the associated variation of protein sequencer are in the COOH-terminal region of ULK3 (NM_001411082.1). To validate the structure predictions, we first aligned them with the crystal structure 4WZX from the Protein Data Bank^40^ using the PyMOL Molecular Graphics System (Version 3.0.4, Schrödinger, LLC).^38^ The 4WZX structure includes a fragment of ULK3 contains 87-amino acids (aa): TSARDLLREMARDKPRLLAALEVASAAMAKEEAAGGEQDALDLYQHSLGELLLLLAAEPPGRRRELLHTEVQNLMARAEYLKEQvkM

For comprehensive evaluation, we predicted the structures of the following three fragments of the COOH-terminal region of ULK3, in response to rs12898397 (T>C on the negative strand), using Google AlphaFold 3.0 for both the VK+ and VK- isoforms:

1) The fragment with the same aa sequence as in the 4WZX crystal structure
2) The 200-aa fragment with an 87-aa region upstream and 26aa downstream of the 4WZX sequence (the 4wzx sequence in uppercases):

ehmpsgeslgratalvvqavkkdqegdsaaalslyckaldffvpalhyevdaqrkeaikakvgqyvsraeelkaivsssnqallrqgTSARDLLREMARDKPRLLAALEVASAAMAKEEAAGGEQDALDLYQHSLGELLLLLAAEPPGRRRELLHTEVQNLMARAEYLKEQVKMresrweadtldkeglsesvrssctlq

3) The fragment with 173-aa, with 5-aa of NH2 terminal and 22-aa COOH terminal being removed from the 200aa-model due to low confidence of their predicted structures

The overlay of these structures was visualized in PyMOL. The structural differences were evaluated using root-mean-square deviation (RMSD) values. An RMSD of 1.0 or lower indicates nearly identical in conformations, while RMSD of 3.0 or higher suggests important structural differences.^41^

SUPPLEMENTAL TABLES

| Supplemental Table 1. GWAS datasets used for MR analysis |
| --- |
| Supplemental Table 2. Sentinel cis-sQTLs in the Framingham Heart Study discovery sample (n=2622) |
| Supplemental Table 3. Sentinel trans-sQTLs in the Framingham Heart Study discovery sample (n=2622) |
| Supplemental Table 4. Replication of the top 10,000 and 100,000 sQTL-isoform pairs from FHS in JHS |
| Supplemental Table 5. Replication of significant cis-sQTLs from FHS in JHS (p<1e-4) from eligible pairs |
| Supplemental Table 6. Enrichment analysis of top cis-SQTLs with GWAS SNPS |
| Supplemental Table 7. Enrichment analysis of top trans-SQTLs with GWAS SNPS |
| Supplemental Table 8. Detected transcripts for OAS1, ULK3, and CNN2 genes |
| Supplemental Table 9. Mendelian randomization analysis of isoforms in three genes to cardiovascular disease traits |
| Supplemental Table 10. Relevant eQTLs and sQTLs in ULK3 gene |
| Supplemental Table 11. Replication of sQTL-sGene pairs in the Garrido-Martin study |

SUPPLEMENTAL FIGURES

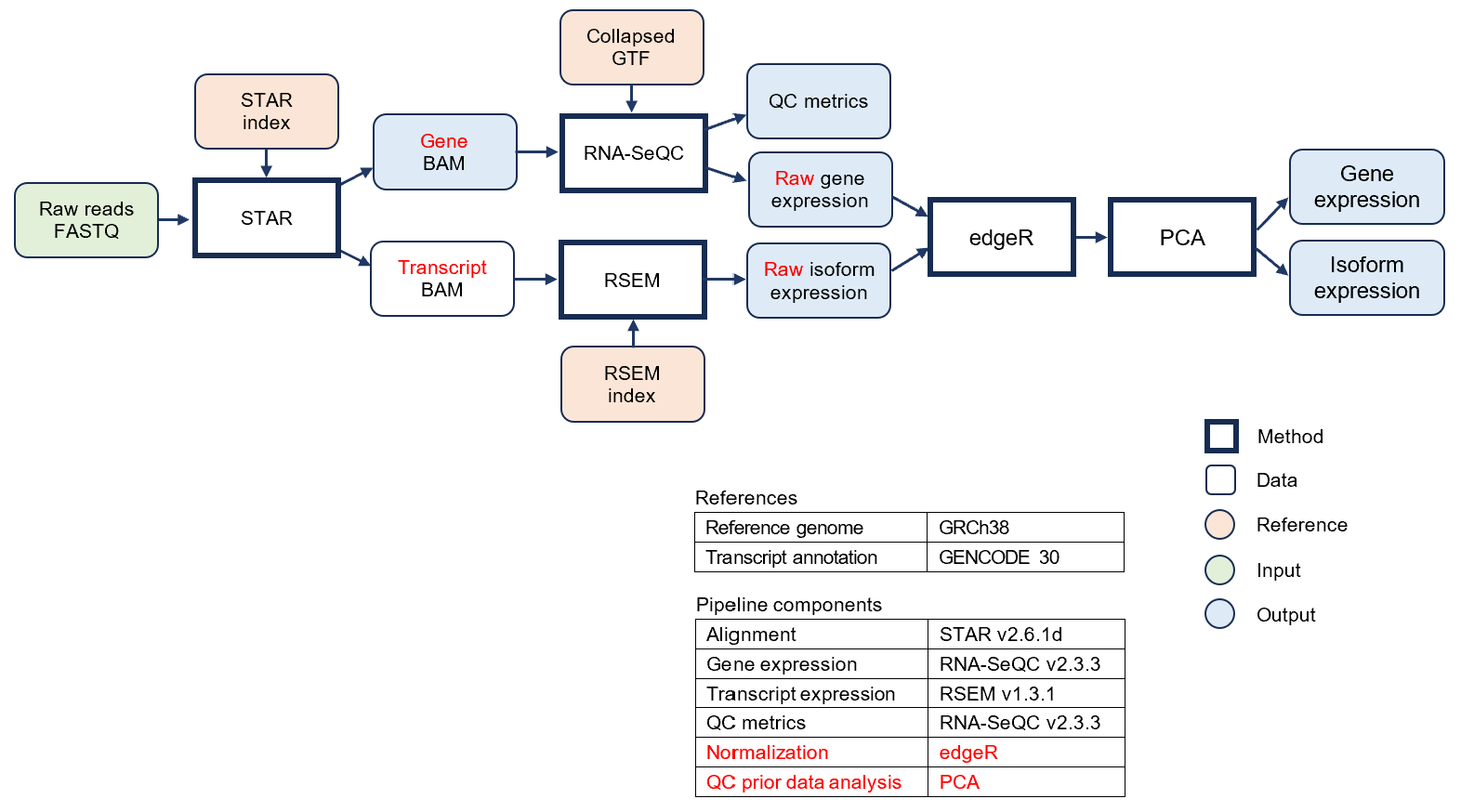

**Supplemental Figure 1**. Flowchart of processing and quality control of RNA-seq data in TOPMed.

https://topmed.nhlbi.nih.gov/sites/default/files/TOPMed_RNAseq_pipeline_flowchart_COREyr3.pd

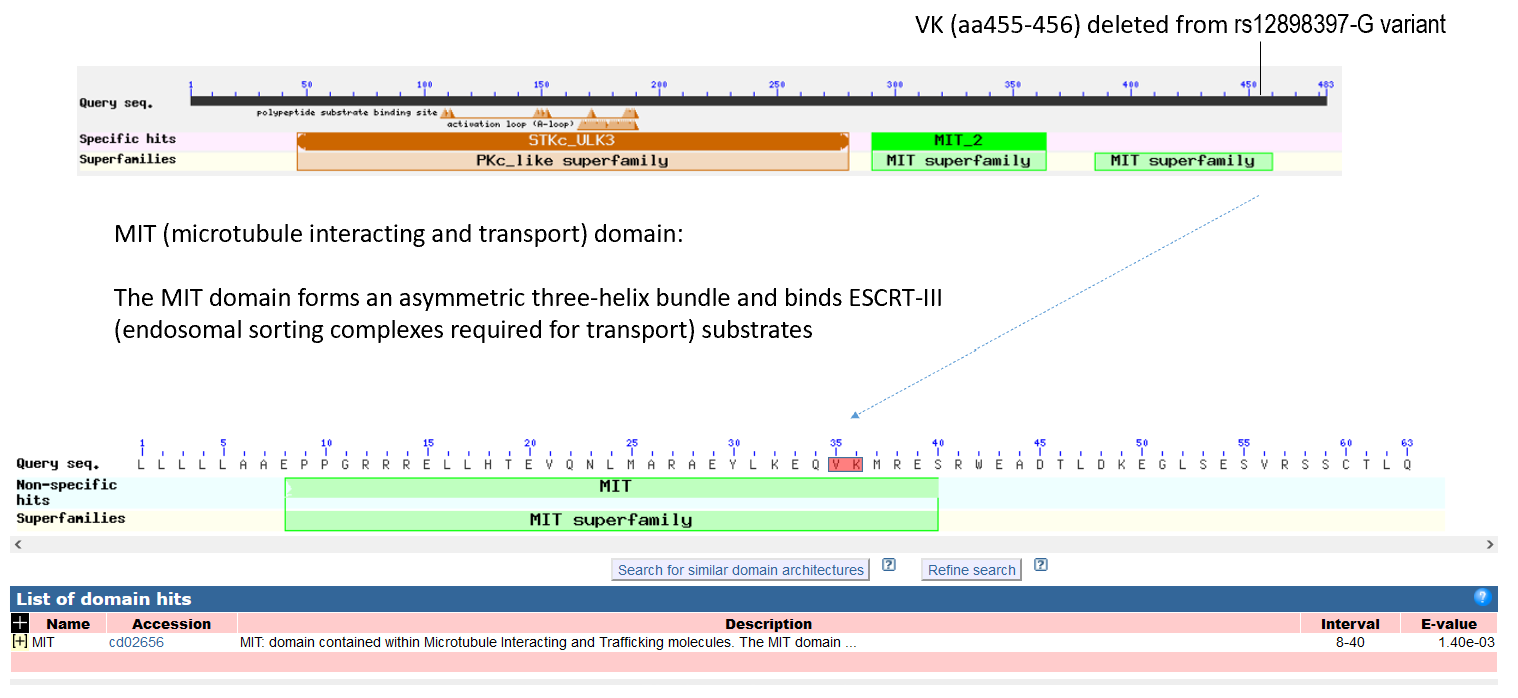

**Supplemental Figure 2**. **rs12898397-G>A region**. MIT, microtubule interacting and transport domain. The rs12898397-G is the sentinel *cis*-sQTL for transcript ENST00000440863.7 (*ULK3*-201) which produces a full-length protein with 472 amino acids, whereas isoform ENST00000569437.5 (*ULK3*-220) produces a truncated protein with two amino acids (“VK”) missing at the 5’ of the 14^th^ exon (**Figure 5A**).

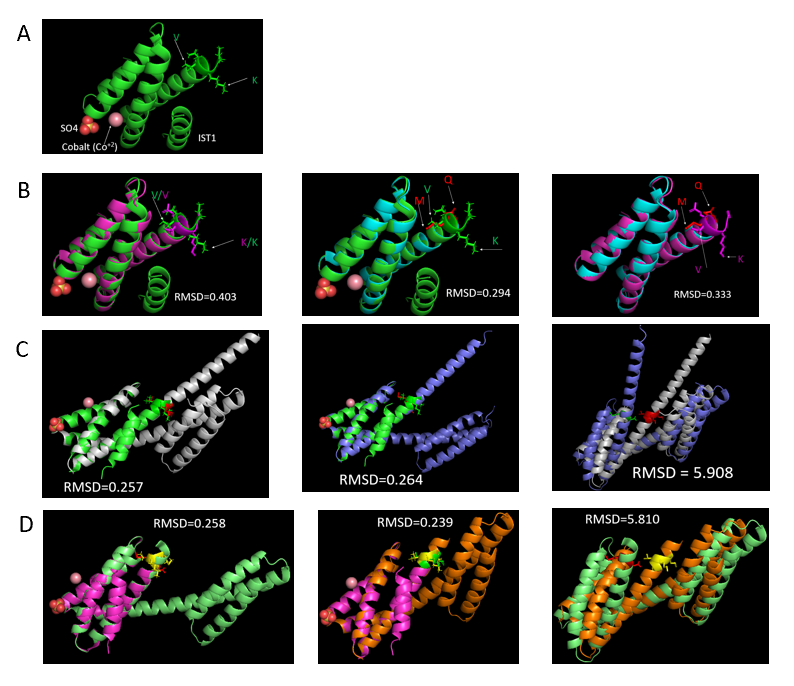

**Supplemental Figure 3**. PyMOL visualization of AlphaFold 3.0-predicted COOH-terminal domains of ULK3 protein variants with (left) and without (middle) "VK" amino acids (aa). The structural differences were evaluated by superimposition (right) using root-mean-square deviation (RMSD) values. An RMSD of 1.0 or lower Indicates nearly identical in conformations, while RMSD of 3.0 or higher suggests significant structural differences

A. The crystal structure of the ULK3 COOH-terminal region, 4WZX (87-aa), complexed with IST1, sulfate, and cobalt ions are shown. The 87-aa sequence is: TSARDLLREMARDKPRLLAALEVASAAMAKEEAAGGEQDALDLYQHSLGELLLLLAAEPPGRRRELLHTEVQNLMARAEYLKEQvkM

B. Alignment of 4WZX with the ULK3 region containing the same aa sequence shows consistent configurations, with RMSD = 0.403 (with “VK”) on the left, and RMSD = 0.294 (without “VK”) in the middle. The structures of the ULK3 region with and without “VK” show similar conformations (RMSD = 0.333).

C. The 87-aa region upstream and 26aa downstream of the 4WZX sequence are added (the 4wzx sequence in capital case):

ehmpsgeslgratalvvqavkkdqegdsaaalslyckaldffvpalhyevdaqrkeaikakvgqyvsraeelkaivsssnqallrqgTSARDLLREMARDKPRLLAALEVASAAMAKEEAAGGEQDALDLYQHSLGELLLLLAAEPPGRRRELLHTEVQNLMARAEYLKEQVKMresrweadtldkeglsesvrssctlq

Alignment of 4WZX with the 200-aa AlphaFold COOH-terminal model, with and without "VK," shows RMSD values of 0.257 (with "VK") and 0.264 (without "VK"). The 200-aa with "VK" and 198-aa without "VK" show different conformations (RMSD = 5.908).

D. The 173-aa model removes 5-aa of the NH2-terminal and 22-aa of the COOH-terminal low-confidence segments. Alignment of 4WZX with the 173-aa AlphaFold model shows RMSD values of 0.258 (with "VK") and 0.239 (without "VK"). The 173-aa model with "VK" and 171-aa without "VK" shows significant conformational differences (RMSD = 5.810). The 5-aa of the NH2-terminal and 22-aa of the COOH-terminal low-confidence segments are shown in **Supplemental Figure 4**.

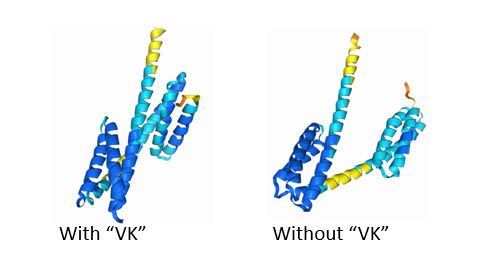

**Supplemental Figure 4**. AF3 prediction of ULK3 200-aa COOH-terminal structure of ULK3 protein with and without “VK”. The 22-aa in COOH-terminal terminal and 5-aa of the NH2-terminal show low confidence in yellow or orange colors. There is also a fragment of low confidence region in the middle of this fragment.
